# Relationship between apnoea duration and changes in physiology in preterm neonates: a systematic review and meta-analysis

**DOI:** 10.1101/2025.04.04.25325244

**Authors:** Yiru Chen, Coen S. Zandvoort, Luke Baxter, Odunayo A. T. Fatunla, Vithushanan Ketheeswaranathan, Ravi Poorun, Zara Small, Fatima Usman, Matthew Henry, Luc Berthouze, Mauricio Villarroel, Caroline Hartley

## Abstract

**Background:** Apnoea is a common respiratory complication in preterm neonates, leading to substantial changes in physiology. We conducted this systematic review and meta-analysis to examine the relationship between apnoea duration and changes in physiology in preterm neonates, and to identify factors that modulate this relationship.

**Methods:** We searched Medline, EMBASE, PsycINFO, and Cochrane Central Register of Controlled Trials databases and included primary empirical studies examining the relationship between apnoea or respiratory pause duration and at least one outcome (heart rate, blood oxygen saturation, cerebral oxygenation, cerebral blood volume) in hospitalised neonates with postmenstrual age (PMA) <37 weeks. Risk of bias was assessed using the Joanna Briggs Institute Critical Appraisal checklist. Results were synthesised narratively and quantitative data was pooled for meta-analysis.

**Results:** Forty-two papers were included, involving a total of 1,483 neonates with 2,399 study sessions. The decrease in heart rate, oxygen saturation, and cerebral oxygenation were significantly correlated with apnoea duration. PMA significantly modulated the relationship, with younger neonates more likely to exhibit oxygen desaturation from short apnoeas.

**Conclusions:** These findings indicate that shorter apnoea alarm thresholds should be considered for younger infants.

Impact statement:

**What is the key message of your article?:**

- Systematic review and meta-analysis of the relationship between change in physiology and apnoea duration in preterm infants.

**What does it add to the existing literature?:**

- Through meta-analysis, we demonstrate that postmenstrual age plays a significant modulating role in the relationship between apnoea duration and change in oxygen saturation, with younger infants more likely to have significant desaturations.

**What is the impact?:**

- We propose that age-stratified apnoea alarm limits are considered to prevent physiological instability in newborns.

## 1. Introduction

Globally, approximately 10% of neonates are born preterm (before 37 weeks of gestation)^1^. Apnoea, the cessation of breathing, is a common respiratory complication for these neonates, affecting more than 50%, especially those born extremely prematurely (before 28 weeks of gestation)^2^. Apnoea can occur multiple times per day and can lead to significant physiological changes, including alterations in heart rate, blood oxygen saturation, cerebral oxygenation, and cerebral blood volume^3–6^. While most episodes resolve spontaneously or with minimal intervention, recurrent or prolonged episodes may have clinical significance and require appropriate monitoring and management. Additionally, apnoea has been linked to long-term complications, such as an increased incidence of retinopathy of prematurity (ROP)^7^ and cognitive deficits later in life^8,9^. These may be related to the extent of apnoea-induced hypoxia or brain activity changes^10^.

There is wide variability within and between neonates in how they respond to an apnoea of a given duration, with short pauses in breathing sometimes leading to large changes in physiology and other times much longer pauses in breathing not leading to significant changes in physiology^4^. Factors such as the age of the neonate may alter physiological responses to an apnoea, with older babies able to tolerate longer periods of apnoea without changes in physiology. It also seems plausible that co-morbidities and medication have an impact. For example, caffeine reduces the incidence of apnoea and intermittent hypoxaemia^11–13^. A better understanding of the factors which modulate the impact of apnoea on physiology is crucial to identify neonates at the highest risk of adverse physiological changes. Ultimately, this could lead to the development of predictive models that can guide treatment or facilitate closer monitoring for high-risk neonates.

We conducted this systematic review and meta-analysis to examine the current knowledge regarding the relationship between apnoea duration and physiological changes (heart rate, oxygen saturation, cerebral oxygenation, and cerebral blood flow) in preterm neonates, and to investigate factors which modulate these relationships. Specifically, we aimed to investigate the following questions:

1. What are the relationships between the duration of pauses in breathing/apnoea and changes in heart rate, blood oxygen saturation, cerebral oxygenation, and cerebral blood volume in hospitalised premature neonates?
2. How do these relationships vary across neonates with different postmenstrual age (PMA), pathology (specifically sepsis and necrotising enterocolitis [NEC]), medication (specifically methylxanthines and opioids), and any other potential modulating factors identified in the included papers?
3. How are these relationships modulated by the frequency/clustering of pauses in breathing?

## 2. Methods

This systematic review is reported in accordance with the Preferred Reporting Items for Systematic Reviews and Meta-Analyses (PRISMA) statement^14^. The protocol for this systematic review was registered in PROSPERO (CRD42024534164).

### 2.1 Eligibility criteria

All primary empirical and peer-reviewed studies reporting a relationship between apnoea or respiratory pause duration and at least one of the outcomes (change in heart rate, blood oxygen saturation, cerebral oxygenation and cerebral blood volume) in hospitalised human neonates with PMA < 37 weeks were included. We excluded papers with the wrong population (non-human species; pre-natal humans; full-term neonates (PMA ≥ 37 weeks) and older (e.g., children, and adult subjects); neonates with neurological abnormalities (e.g. seizures) or congenital abnormalities; and non-hospitalised neonates. We also excluded papers with the incorrect study characteristics (secondary literature (e.g. reviews, book chapters); non-empirical literature (e.g. opinions, commentaries, perspectives); non-peer-reviewed grey literature (e.g. conference abstracts, meeting reports, theses); and study protocols). There were no restrictions based on publication language or date.

### 2.2 Search strategy

A combination of subject headings terms and controlled keywords were used, and searches were conducted initially on 23 November 2023 and updated on 10 December 2024 in the following databases: MEDLINE (Ovid), Embase (Ovid), PsycINFO (Ovid) and Cochrane Central Register of Controlled Trials (Cochrane Library, Wiley). Detailed searching strategies are included in Supplementary Methods. Additionally, backward citation tracking was carried out on the final set of papers as an additional information source, and this was done through the Citationchaser package^15^.

### 2.3 Selection process

A systematic review management tool, Covidence (Melbourne, Australia), was used to manage records and data throughout the review. The references were imported onto the platform, and automatic de-duplication was performed. In total, seven reviewers were involved in the selection process. The selection followed a two-stage process:

1. A title and abstract screening against the inclusion criteria to identify potentially relevant papers.

2. A full-text screening of all the papers identified as possibly relevant for inclusion from the initial screening above.

At the title and abstract screening stage, a pilot block of randomly selected 20 articles was screened by all reviewers to ensure consistency of reviewers across the screening process. Papers were then divided into blocks, with two independent reviewers assigned to each block. Papers whose abstract did not describe any form of relationship between apnoea and changes in vital signs were excluded. At the full-text screening stage, the same two reviewers assessed all papers, except those written in non-English languages. Papers written in Dutch, German, Italian, or French were reviewed by fluent speakers of the respective language, with an additional reviewer using Google Translate to validate the results. One paper in Bulgarian and one in Danish were screened using Google Translate only and excluded at the title and abstract screening stage. At both stages, screening was carried out in duplicate and independently by the two reviewers, and inconsistencies were settled by discussion between both reviewers and if necessary, an arbitrator. For consistency, one reviewer (YC) acted across all papers. The same arbitrator (LBa) also acted across all papers.

### 2.4 Data collection process and data items

Data extraction was carried out by a single reviewer (YC). When relevant information was provided only in figure format, the PlotDigitizer software was used to extract values from the figures. This process was conducted twice for each figure to ensure consistency in the results. The following variables were extracted: author’s names; year of publication; country and region of publication (according to World Health Organisation categorisation^16^); hospital(s) where data was collected, study period; study design; sample size; neonate’s age (gestational age and postmenstrual age), including median and range where available; type of respiratory support received by the neonates (if any); the pathological conditions of the neonates (specifically sepsis and NEC); type of medication used (if any); description of apnoea/pause in breathing (i.e., duration, method of measurement, any resuscitation provided, information on the frequency of pause/clustering of pauses); description of the changes in outcome variables (i.e., absolute change, baseline and minimum/maximum values if provided, method of measurement); duration of the recordings and time of day; signal quality measures (if given).

### 2.5 Study risk of bias assessment (quality assessment)

Assessment of the risk of bias in the included studies was carried out by using the checklist for systematic reviews and research syntheses provided by the Joanna Briggs Institute Critical Appraisal tools. Two reviewers (YC and CZ) evaluated the risk of bias independently. Any disagreement between the two reviewers was settled through discussion.

To derive an overall risk of bias rating^17,18^, we focused exclusively on items within the JBI checklists related to internal validity. Each item in the relevant JBI checklists was classified as either “critical” (pertaining to internal validity) or “non-critical” (pertaining to other constructs). Critical items were mapped to established domains of bias, including selection bias, confounding, measurement bias, temporal bias, and attrition bias. Studies were then rated based on their responses to critical items, following a predefined rule set: low risk of bias was defined as no “no” responses on critical items; moderate risk of bias was defined as one “no” response on a critical item; high risk of bias was defined as two or more “no” responses on critical items. Responses of “unclear” or “not applicable” were not treated as indicators of bias, ensuring a conservative and transparent assessment approach.

### 2.6 Data synthesis

The outcome of the database searches and study selection process was presented in a PRISMA flowchart. The data synthesis was performed using a comprehensive narrative approach to ensure a rigorous and informative analysis. Narrative and qualitative synthesis involved the tabulation of results where feasible.

Quantitative data from the selected studies regarding the relationship between apnoea duration and physiological changes in preterm neonates were extracted and pooled for meta-analysis where available. The dependent variable were the mean percentage changes in vital signs. If a study provided the values for percentage change in vital signs directly, these were used for the meta-analysis. When values at the beginning and end of apnoea were provided, the difference was calculated and divided by the starting value to determine the percentage change. The independent variable was the apnoea durations. If a study provided the mean apnoea durations for each pooled point, these were used directly. When only a range of apnoea durations was provided, the midpoint of the range was used as the X-value (e.g., if the range of apnoea duration was 10 – 20 seconds, the X-value would be 15 seconds for that point). ^19,20^ If multiple studies presented the same results using the same dataset, then we only included the results once in the meta-analysis.

Pooled data were analysed using Pearson correlation coefficients, linear regression models, and linear mixed-effects models. For the linear regression models, apnoea duration was the predictor, and changes in vital signs were the response. The square root of the number of apnoea events associated with each data point was used as the weight in the model. The coefficient of determination (R^2^) was used to assess how much of the variation in vital signs could be explained by apnoea duration. The formula to calculate R^2^ was:

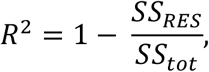

where *SS_RES_* is the sum of squares of residuals, and *SS_tot_* is the total sum of squares.

For the linear mixed-effects models, the study ID number was used as a random effect, and apnoea duration as a fixed effect. The impact of apnoea type, PMA, and the use of theophylline (a medication that is commonly used for apnoea treatment) on the relationship between apnoea duration and physiological changes was analysed, where applicable, by adding interaction terms between these factors and apnoea duration as fixed effects in the model.

For a post hoc analysis of how PMA affects the relationship between changes in oxygen saturation (SpO2) and apnoea duration, we used a linear regression model with the formula:

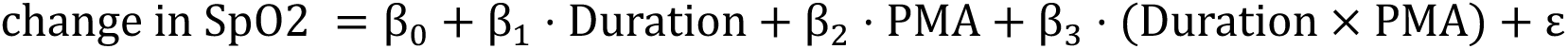

Using this model, we predicted the apnoea duration at which oxygen saturation is likely to drop by more than 10% for neonates of different PMAs. The 10% threshold is commonly used for defining neonatal desaturation^21^.

The correlation coefficients from different papers reporting the same outcome were standardised and converted into a common effect size metric, Fisher’s Z score, which was combined to provide overall correlation factors. The regression lines from the individual studies were plotted alongside the line obtained from the meta-analysis for visual comparison of the variation across studies.

## 3. Results

A total of 4,362 studies were identified, after the removal of 8,567 duplicates (Figure 1). Following the title and abstract screening, 4,190 studies were excluded, and a further 129 studies were excluded after the full-text review. One study^22^ was excluded due to unavailability of the full text after database searches and inter-library requests. Overall, 42 studies were included in this review, involving a total of 1,483 neonates with 2,399 recording sessions (i.e. a period during which a patient’s vital signs are recorded). The median number of neonates in each study was 24 (range: 1 – 335, IQR: 14 – 31.75), and the median number of recording sessions was 31 (range: 1 – 386, IQR: 18 – 78). Twenty-three of the included studies (54.8%) were cross-sectional, 16 (38.1%) were cohort studies, 2 (4.8%) were case-control studies, and 1 (2.4%) was a case report. The year of publication ranged from 1969 to 2022, with a median of 1992. Most of the studies were from Europe (n = 21, 50%) and the Americas (n = 17, 40.5%). The remainder of studies were from the Western Pacific (n = 4, 9.5%). No studies were published from the South-East Asian Region, Eastern Mediterranean or Africa (regions classified using the World Health Organisation categorisation^23^).

**Figure 1:**
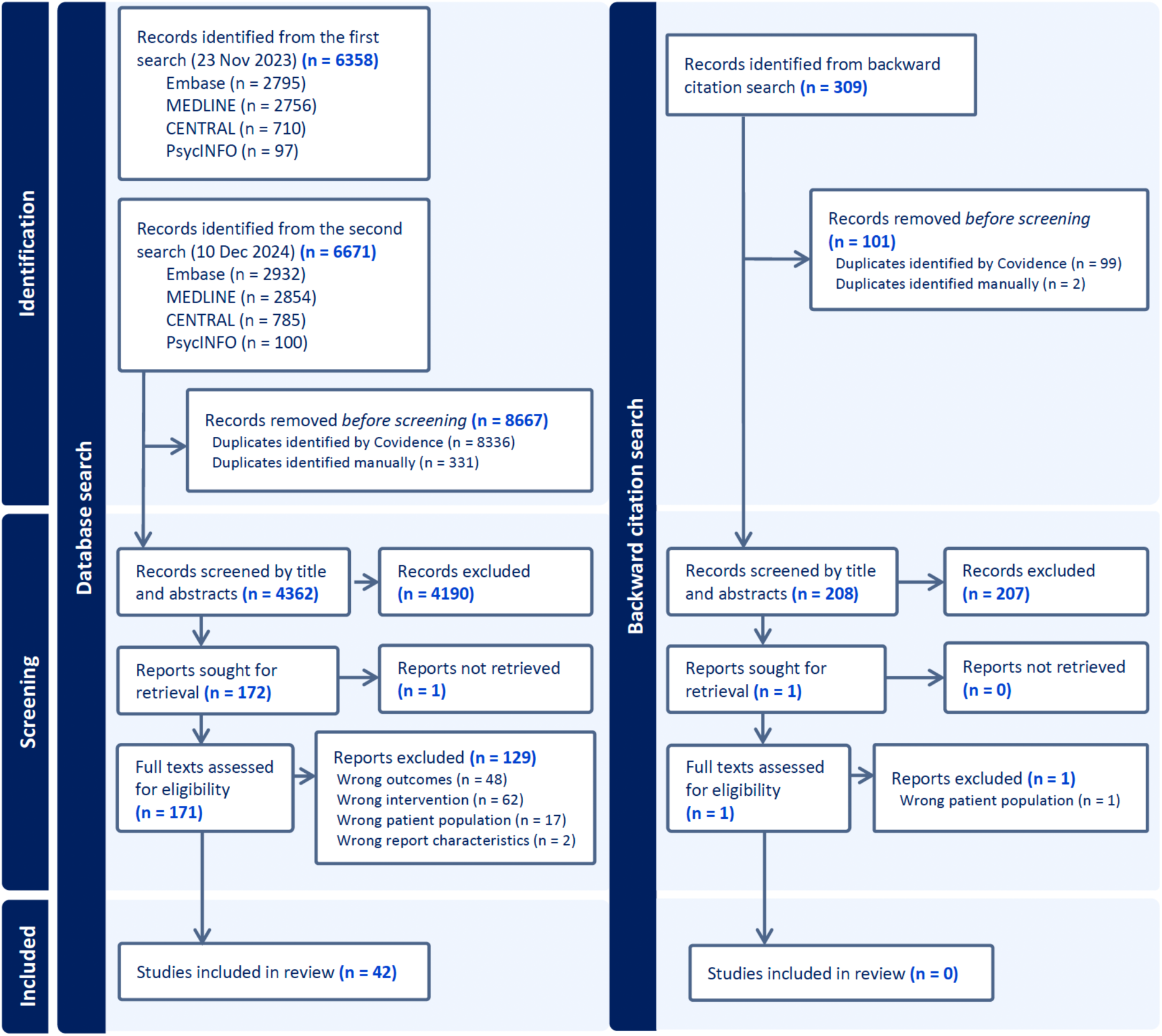
Prisma flow chart showing the study selection process.

Thirty-one articles (73.8%) described the relationship between apnoea duration and change in heart rate, 24 articles (57.1%) described the relationship between apnoea duration and change in oxygen saturation, 3 studies (7.1%) described the relationship between apnoea duration and change in cerebral oxygenation, and only one study (2.4%) described the relationship between apnoea duration and change in cerebral blood volume.

The definition of apnoea and pauses in breathing investigated in the different studies varied; some studies included pauses in breathing as short as 3 seconds, while other studies only included apnoeas with a duration of 20 seconds or more. The techniques used to measure apnoea also varied, including polysomnography, acoustic monitoring, inductive plethysmography, thermistor, and pneumotachograph (Tables 1-4).

**Table 1:** Summary of included studies reporting the relationship between apnoea duration and change in heart rate. ECG: Electrocardiogram. SD: standard deviation.

| Study | Number of infants | Postmenstrual age at study (weeks) | Definition or inclusion criteria of apnoea/pause in breathing | Method of apnoea/pause measurement | Description of apnoea/pause in breathing | Method of outcome measurement | Main results | Quality assessment |
| --- | --- | --- | --- | --- | --- | --- | --- | --- |
| Daily et al. 1969<sup>3</sup> | 22 | Premature | Apnoeic episodes of 20 or more seconds. | Impedance plethysmography (impedance pneumography) | A total of 540 apnoeic episodes were observed in 13 infants. 400 episodes occurred in 8 of 14 infants who were otherwise clinically well. 29 episodes occurred in 4 of the 7 infants recovering from respiratory distress syndrome. Of the 540 episodes, 428 exceeded 30 seconds in duration. | ECG | For apnoea > 45 seconds, heart rate fell steadily; in episodes of shorter duration, there was a tendency for the heart rate to rise in the first 5 seconds and thereafter to fall. | Inclusion criteria was not stated. No confounding factors identified. No statistical analysis performed. |
| Gabriel et al. 1976 (A)<sup>17</sup> | 8 | Range: 31 - 35. | Respiratory arrest: pauses > 3 seconds. Apnoeic spells: respiratory arrest of more than 10 seconds duration. | Polysomnography | 493 apnoeic spells and 2450 respiratory pauses. | ECG | Pauses < 10 s: no correlation between the duration of the respiratory arrest and the incidence of bradycardia. Pauses ≥ 10 seconds: the incidence of cardiac slowing as well as the percentage of bradycardia increased with longer duration of apnoeas. | Method of exposure and outcome measurements were unclear. No statistical analysis performed. |
| Gabriel et al. 1976 (B)<sup>18</sup> | 8 | Range: 31 - 35. | Breathing pauses are categorised into two groups: pauses lasting 3–10 seconds and apnoeas which exceeded 10 seconds. | Polysomnography | n = 2,450 pauses of 3-10 seconds, n = 493 apnoeas | ECG | Most of the breathing pauses (especially apnoeas) are accompanied by bradycardia. The number of bradycardias increased with increasing apnoea duration. | Method of exposure and outcome measurements were unclear. No statistical analysis performed. |
| Storrs 1977<sup>43</sup> | 9 | Range: 27.3 – 34.4. | Apnoea: cessation of spontaneous respiratory effort for > 5 seconds coinciding with cardiovascular changes. | Plethysmography | Total of 47 apnoeas | ECG | The heart rate fell by a mean (±SD) of 43% ± 13%, on average 8.6 seconds ± 6.3 (range 2-33 s) after the onset of apnoea, rising on average 4.8 seconds ± 4.4 (range 2-16 s) after the resumption of breathing. The maximum fall in heart rate is correlated to the apnoea duration. | Method of exposure measurement was unclear. Statistical analysis method was also unclear. |
| Fenichel et al. 1980<sup>26</sup> | 11 newborns, only 6 newborns eligible for this review, the remaining 5 had conceptional age $\geq 37$ weeks. | 32; 32; 36; 33; 33; 35. | Not given. | Thermocouple or piezoelectric transducer. | A total of 16 nonconvulsive apnoeic spells were recorded in the eligible patients. Nine of them were between 4-19 seconds, and 7 were $\geq 20$ seconds. | ECG | Alterations in heart rate differed between the two groups (apnoea duration $< 20$ s and $\geq 20$ s). During apnoea lasting less than 20 seconds, heart rate was found to increase, decrease, or remain unchanged. During apnoea lasting 20 seconds or longer, heart rate usually fell by 40% or more within 8 seconds of the onset of apnoea. | The inclusion criteria and the population details were not clearly stated. The measurement of exposure was unclear. The measurement of outcome was unreliable. Confounding factors were not identified. No statistical analysis performed. |
| Vyas et al. 1981<sup>30</sup> | 7 | Mean: 34.1. | Apnoea: cessation of tidal flow for $> 5$ seconds. | Pneumotachograph | Total of 172 episodes of apnoea.<br>Number (percentage): obstructive - 86 (50%); central - 48 (27.9%); mixed - 38 (22.1%).<br>Mean apnoea duration (sec): obstructive - 9.93; central - 8.3; mixed - 10.9. | ECG | The lowest heart rate, the mean duration of heart rate below 100 bpm, and the incidences of bradycardia all differed among the three apnoea type groups. Central apnoea tended to have the shortest duration and the smallest changes in heart rate, whereas obstructive and mixed apnoeas were associated with longer durations and greater changes in heart rate. | No statistical analysis performed. |
| Werthamm<br>er et al.<br>1983<sup>44</sup> | 8 | Premature. | Apnoea: cessation of respiration for > 15 seconds associated with a decrease in heart rate. Classed as mild when heart rate decreased by 20 beats per minute below the average of the previous two minutes, and as severe when the heart rate fell below 100 beats per minute. | Acoustic monitoring | Total of 26 apnoeas. 17 mild, 9 severe.<br>Cessation of breathing (seconds) for the 17 mild episodes: mean 18.2, SD 4.3, range 15-30. Information not given for severe episodes. | ECG | During apnoeic spells that became severe, a mean 27.5 ± 9.7 (SD) seconds elapsed after cessation of respiration before the heart rate fell to less than 100 beats per minute, which is longer than the average duration of mild episodes. | The inclusion criteria and the population details were not clearly stated. Confounding factors were not identified. No statistical analysis performed. |
| Southall et al. 1983<sup>34</sup> | 14 (13 eligible for this review, subject 10 excluded due to infant's age) | Mean: 31.8, range: 28 - 25. | Apnoeic episodes ≥ 20 seconds and ≥ 30 seconds. | Impedance respirometer, under-mattress pressure pad, segmented air-filled mattress, pressure capsule system. | A total of 203 apnoeic episodes ≥ 20 seconds, and 120 episodes ≥ 30 seconds were identified. Duration of apnoea ≥ 30 seconds (s): mean: 46, range: 30 - 216. | ECG | Of a total of 203 episodes of apnoea, 145 (71%) were accompanied by bradycardia of 80 bpm. Duration of apnoea is correlated with the duration of accompanying bradycardia. | Confounding factors were not identified. No statistical analysis performed. |
| Henderson-Smart et al. 1986<sup>31</sup> | 28 | Range 28 - 44. | All events of 10 seconds or more in duration were noted. | Mercury in rubber strain gauge, thermistor. | There were 1520 apnoeas of over 10 seconds in duration, 1002 with a duration of 10-14 seconds, 311 with duration 15-20 seconds and 207 of more than 20 seconds. | ECG | The major factor influencing the incidence of bradycardia was duration of apnoea. For each apnoea type there was a significant increase in the incidence of bradycardia as duration of apnoea increased (p<0.001 in each case). Time to the onset of the fall in oxygen saturation and to the onset of bradycardia became longer with increasing apnoea duration. | The strategies to deal with confounding factors were not stated. |
| Muttitt et al. 1988<sup>21</sup> | 27 | Preterm. | The apnoea alarm was set to 15 seconds. This alarm notifies staff of a cessation of respiratory input after 10 seconds by way of an intermittent beep which turns into a continuous alarm after 15 seconds. | Impedance respirometry | A total of 1266 apnoeas were documented in the 27 infants. 28% of the apnoeas were less than 20 seconds in duration, 48% were between 20 and 30 seconds, 21% were between 30 and 50 seconds, and 3% were greater than 50 seconds. The longest lasting recorded apnoea was 104.1 seconds. | ECG | A significant correlation was found between apnoea duration and decrease in heart rate ($r = .093$, $P = .001$). | The inclusion criteria and the population details were not stated. The measurement of outcome was unclear. |
| Mathew 1988<sup>32</sup> | 24 | Mean: 35.0, standard deviation: 2.0. | Apnoea: the absence of nasal airflow. Prolonged apnoea: apnoea of 20 seconds or longer duration. Short apnoea: apnoea between 10-19 seconds. | Plethysmography | No episodes of apnoea longer than 10 seconds or bradycardia occurred in 9 infants. 39 episodes of apnoea were observed in the remaining 15 infants. 36 episodes were short, 3 prolonged. | ECG | 9/36 short apnoeas and 2/3 prolonged apnoeas were associated with bradycardia. Prolonged apnoeas are more likely to be accompanied by bradycardia. | The strategies to deal with confounding factors were not stated. The strategies to address incomplete follow up were not utilised. |
| Curzi-Dascalova et al. 1989<sup>46</sup> | 23 infants recruited: 7 eligible for this review, 16 excluded as had post-conceptional age > 37 weeks). | 35-36 week subgroup included in this review | Apnoea: central respiratory pauses > 3 seconds. | Polygraphic recordings | 602 pauses in total. None of them lasted more than 12 s. | ECG | Pauses lasting 10 to 12 seconds were relatively rare compared to shorter pauses and did not result in greater cardiac deceleration than shorter pauses. | The inclusion criteria and the population details were not stated. The population details and study settings were unclear. The measurement of exposure was unclear. No statistical analysis performed. |
| Suichies et al. 1989<sup>22</sup> | 18 | Preterm | Apnoeic spell: the cessation of breathing for a minimum of 6 seconds. | Thoracic impedance, thermistor | A total of 212 apnoeic spells were analysed. The mean duration of an apnoeic spell was 10.3, 10.2 and 14.3 s for central, obstructive and mixed apnoeic spells, respectively. | ECG | A positive correlation was observed between apnoea duration and the percentage of episodes accompanied by bradycardia. The correlation coefficients varied by apnoea type: central apnoea ($r = 0.65$, $p < 0.01$), obstructive apnoea ($r =$ | The inclusion criteria and the population details were not stated. The population details and study settings were unclear. The measurement of exposure was unclear. |
| | | | | | | | 0.51), and mixed apnoea ($r = 0.56$, $p < 0.05$). | Confounding factors were not identified. |
| Waggener et al. 1989<sup>35</sup> | 14 | Premature | Apnoea: cessation of airflow at mask for a minimum of 3 seconds. | Pneumotachograph, magnetometers. | 182 apnoeas $\geq 3$ s in duration, 38 apnoeas $\geq 10$ in duration. | ECG | The decrease in heart rate during apnoea was modelled as a function of apnoea duration using multiple linear regression and dichotomous logistic regression. The contingency table based on logistic regression showed that 87 out of 99 events with a decrease of $<10$ bpm and 24 out of 41 events with a decrease of $\geq 10$ bpm were successfully predicted, resulting in an average accuracy of 73%. | Confounding factors were not identified. No statistical analysis performed. |
| Hodgman et al. 1990<sup>36</sup> | 66 | Range: 32 - 36. | The duration of an apnoeic episode was measured from the end of the inspiration before the pause to the beginning of the next inspiration. | Impedance pneumography. | Approximately half of the infants (53%) had no apnoeic episodes of 15 seconds or longer. 25 (38%) infants had one or more 15- to 19-second apnoea, and in 10 infants (15%), the duration was 20 seconds or longer. Four of these 10 infants (6% of the total population) had two or more episodes of apnoea of this duration.<br>There was a total of 94 individual apnoeic episodes of 15 seconds or longer. Twenty of the 94 episodes lasted 20 seconds or longer, and 5 of these were between 30 and 40 seconds in duration. | ECG | A TEB (transient episodes of bradycardia) was defined as a drop in heart rate below 90 beats per minute. While the majority of TEB did not accompany a long apnoeic episode, all but 5 of the apnoeic episodes of 15 seconds or longer were accompanied by a TEB. | The measurement of exposure was unclear. Confounding factors were not identified. Methods of statistical analysis were unclear. |
| Mathew et al. 1991<sup>37</sup> | 10 | Preterm. | Apnoeic episodes: absence of airflow for 5 seconds or greater.<br>Apnoeas lasting $\geq 15$ seconds were termed long apnoeas, whereas those lasting $< 15$ seconds were termed short apnoeas. | Plethysmography, nasal Screen Flowmeter | A total of 352 apnoeic events. Out of the 202 episodes preceded by arousal, most of the apnoeas were short: 181 were less than 15 s, and only 21 were $> 15$ s. | ECG | Hypoxia or bradycardia developed during 24% of long apnoea ($\geq 15$ s) and only 7% of short apnoea ($< 15$ s). | The inclusion criteria and the population details were not stated. Confounding factors were not identified. No statistical analysis performed. |
| Upton et al. 1991<sup>4</sup> | 27 | Median: 32, range: 26 - 36. | Apnoea: cessation of breathing movement for at least 10 seconds, or a shorter period if associated with bradycardia of 90 beats per minute or less. | Thoracic impedance, plethysmography. | A total of 1029 episodes were analysed.<br>Duration of apnoea (s): median 10 (range 0-80) for infant receiving theophylline (45 studies, 535 episodes); median 11 (range 0-31) for infants not receiving theophylline (44 studies, 494 episodes). | ECG | Correlation between minimum heart rate during episodes of bradycardia and apnoea is extremely poor. Minimal heart rate=74.22-0.14*duration of apnoea ($r=0.11$, $p<0.0001$). | The measurement of exposure was unclear. The strategies to deal with confounding factors were not stated. |
| Spear et al. 1992<sup>38</sup> | 26 | All patients - mean: 34, SD: 2.<br>Patients with prolonged apnoeas - mean 33, SD: 1.<br>Patients with no prolonged apnoeas - mean: 35, SD: 2.<br>p = 0.001. | Prolonged apnoea: > 20 seconds, short apnoea: 6 - 20 seconds. | Thoracic impedance, nasal air flow (by thermistor). | Of the 78 traces, 20 had prolonged apnoea and 58 had no prolonged apnoea.<br>Traces had a mean of 16.1 episodes of central apnoea, 3.04 episodes of obstructive apnoea, and 6.1% of periodic breathing. | ECG | The number of bradycardias > 5 seconds were greater in the traces with prolonged apnoea comparing to short apnoea. | The two groups were not similar in study weight, post-conceptional age and postnatal age. No statistical analysis performed. |
| Finer et al. 1992<sup>27</sup> | 47 | Premature | Apnoea was included in this study if it lasted at least 15 seconds and was associated with bradycardia, hypoxaemia, or both. | Impedance respirometry | 2082 episodes of apnoeas in total. | ECG | As apnoea duration increased, the percentage fall in heart rate decreased, this trend being significantly greater for central and obstructive apnoea than for mixed events. As the duration of central apnoea became longer, untreated infants had a significantly lesser fall in heart rate; | The inclusion criteria and the population details were not stated. The details of the study subjects and the setting were unclear (study period was not stated). The measurement of exposure was unclear. |
| | | | | | | | apnoea between 40 and 50 seconds in duration was associated with a mean fall in heart rate of 38%, compared with 51% during apnoea lasting 20 to 30 seconds ($p = <0.0001$). | Confounding factors were not identified. |
| Upton et al. 1992 (A)<sup>23</sup> | 27 (24 have available flow recordings). | Median: 32, range: 26 - 36. | Apnoeic episodes of at least 5 seconds duration, as detected on the flow trace, were analysed for comparison. | Thoracic impedance, plethysmography. | From the initial polygraphic recording: 86% of the bradycardic episodes were associated with apnoea.<br>During airway flow measurements: apnoea was present on the flow trace in all 41 episodes in which bradycardia occurred. Median apnoea duration was 14 seconds (range 2-51). | ECG | The duration of bradycardia, expressed by the number of bradycardic beats, was positively correlated with apnoea duration in seconds. There was also a significant positive correlation between apnoea duration and time of onset of bradycardia. | The measurements of exposure and outcome were unclear. |
| Poets et al. 1993<sup>42</sup> | 80 | Mean: 36.3, SD: 2.3. | Apnoeic pauses of 4 seconds or longer associated with bradycardia. An association was considered present if commenced within $\pm 15$ seconds of the onset of the bradycardia. | Volume expansion capsule, plethysmography. | 143 (83%) of the 172 bradycardias associated with interpretable airflow and breathing movement signals were accompanied by an apnoeic pause. | ECG | The apnoeic pauses associated with bradycardia had a median duration of 10 seconds, with range between 4 and 38 seconds. | The inclusion criteria and the population details were not stated. The details of the study subjects and the setting were unclear (study period was not stated). The measurements of exposure were unclear. Confounding factors were not identified. No statistical analysis performed. |
| Jenni et al.1996<sup>24</sup> | 17 | Preterm | Apnoea: cessation of respiratory effort for 10 seconds or more. | Thoracic impedance, thermistor. | A total of 130 apnoeic episodes were recorded in 14 infants. The median duration of those episodes was 20 s, the 5th and 95th percentiles were 12 and 54, respectively. Each infant had between 4 and 20 episodes of apnoea (mean 9.3) per study period). | ECG | The duration of apnoea was correlated with the lowest heart rate: central apnoea: $r = -0.772$ ($p < 0.01$), obstructive apnoea: $r = 0.104$, and mixed apnoea: $r = -0.695$ ($p < 0.01$). | The inclusion criteria and the population details were not stated. The measurements of exposure were unclear. Confounding factors were not identified. |
| Carbone et al. 1999<sup>28</sup> | 90 infants recruited, multichannel recording data were complete in 32 infants. | Mean: 36.9. | Episodes of apnoea of $\geq 10$ seconds in duration. | Thermistor. | 236 episodes of apnoea analysed. The long apnoea group consisted of 14 apnoeic events of $\geq 20$ seconds in duration, the medium apnoea group consisted of 50 apnoeic events of 15 to 19 seconds in duration, and the short apnoea group consisted of 172 apnoeic events of 10 to 14 seconds in duration. | Monitor | Longer apnoeas were significantly correlated with a lower heart rate nadir, a longer duration of heart rate below two-thirds of the baseline, a greater drop in heart rate, and a faster rate of heart rate decline. | The inclusion criteria and the population details were not stated. The details of the study subjects and the setting were unclear (study period was not stated). The measurements of exposure were unclear. Confounding factors were not identified. Methods of statistical analysis were unclear. |
| Curzi-Dascalova et al. 2000<sup>41</sup> | 5 | Mean: 34, range: 33.6 - 34.7. | Apnoea: a 20% or greater decrease in the amplitude of abdominal respiratory movements (detected 2 cm above the umbilicus) or/and airflow. | Thermistors. | 492 apnoeas longer than 5 s were detected, of those only 27 were longer than 10 s. | ECG | Multivariate linear analysis showed that the normalised percentage change in HR was dependent on apnoea duration ($p < 0.0001$); regression analysis models explained 24.7% of the changes found, and coefficient of correlation was $r = 0.50$. | The details of the study subjects and the setting were unclear (study period was not stated). The measurements of exposure were unclear. |
| Pichler et al. 2003<sup>5</sup> | Bradycardia group: $n = 20$; Non-Bradycardia group: $n = 19$. | Bradycardia group - mean: 34.2, SD: 2.8.<br>Non-Bradycardia group - mean: 35.0, SD: 2.4.<br>$p$ value $> 0.05$ | Episodes of apnoea lasting longer than 15 s with bradycardia (bradycardia group) were analysed and compared to episodes of apnoea lasting longer than 15 s without bradycardia (non-bradycardia group). | Piezo-respiratory effort sensors | Twenty-six episodes of apnoea with bradycardia and 26 without bradycardia were observed. The mean duration of apnoea episodes was $25.9 \pm 6.0$ s in the bradycardia group and $25.0 \pm 8.6$ s in the non-bradycardia group ($p > 0.05$). | ECG | Longer apnoea durations are associated with greater drops in heart rate in both the bradycardia and non-bradycardia groups. | Unclear whether the two groups were similar and recruited from the same population. Confounding factors were not identified. |
| Nock et al. 2004<sup>39</sup> | 17 | Mean: 35, SD: 1. | Number of apnoeas $\geq 15$ seconds and $\geq 20$ seconds were recorded. Those lasting $\geq 20$ seconds are a subset of the episodes $\geq 15$ seconds in length. | Plethysmography. | The subjects had 0 to 46 apnoeic episodes $\geq 15$ seconds and 0 to 14 episodes $\geq 20$ seconds. | ECG | Heart rate was $119 \pm 17$ (94-163) bpm during apnoeas $\geq 15$ seconds and $110 \pm 11$ (91-126) bpm during apnoeas $\geq 20$ seconds. | The details of the study subjects and the setting were unclear (study period was not stated). The measurements of exposure were unclear. Confounding factors were not identified. |
| Beck et al. 2011<sup>29</sup> | 10 | Mean: 32.1, range: 29.6 - 36.4. | Central apnoeas: periods where the electrical activity of the diaphragm was a flat tracing for more than 5 seconds. | Oesophageal electromyography . | 188 central apnoeas identified. Apnoeas were classified by their duration (5 - 10 s, 10 - 15 s, and > 15 s). | Not given | Longer apnoea durations were associated with greater drops in heart rate during the events. | The details of the study subjects and the setting were unclear (study period was not stated). The strategies to deal with confounding factors were not stated. |
| Mohr et al. 2015<sup>25</sup> | 335 | Preterm | ABD: Apnoea events accompanied by bradycardia (heart rate <100 bpm) and oxygen desaturation (oxygen saturation <80%). ABD events with apnoea lasting more than 10 seconds were recorded and categorised by duration.<br>Very long apnoea (VLA) was defined as breathing cessation >60 seconds with bradycardia and oxygen desaturation. | Chest impedance-derived respiratory rate by central network server. | 86 VLA occurred in 29 infants. The number of events per infant ranged from 1 to 19.<br>In all infants there were 100 events of a duration 10-20 seconds, 100 events of 30-40 seconds, 46 events of 60-80 seconds, and 40 events of 80 seconds. | ECG | The areas under the HR curves increase approximately linearly with the duration of the apnoea. The sample correlation coefficient is 0.81 for heart rate ($p < 0.001$). | The inclusion criteria and the population details were not stated. Confounding factors were not identified. |
| Marshall et al. 2019<sup>33</sup> | 30 | Median: 30, inter-quartile range: 28-33. | Individual respiratory pauses between 5 and 29 seconds (inclusive) were identified. Events with an FiO2 (fraction of inspired oxygen) adjustment (defined as a change in FiO2 of at least 0.01) occurring during the event window were excluded. | Abdominal capsule. | In total, 17677 isolated and 9623 clustered respiratory pause events were identified from a total of 55767 respiratory pauses greater than or equal to 5 seconds. After exclusion of 572 isolated and 443 clustered events because of FiO2 adjustments within the event window, there were 17105 isolated and 9180 clustered events available for analysis. | Monitor | The mean trajectories of heart rate over the event window demonstrate more prominent instability after longer duration pauses, both for pauses that were isolated and clustered. The proportion of events followed by physiological instability increased with longer pause duration, and was higher for clustered events for all pause durations. | The exposure and outcomes were measured in valid and reliable ways. Confounding factor (clustering of apnoea) and the strategy to deal with the factor were stated. |
| Falsaperla et al. 2019<sup>45</sup> | 1 | 29.6 | Apnoea: cessation of airflow lasting more than 20 seconds, or cessation of airflow associated with bradycardia (20% below the baseline heart rate), or cessation of airflow for less than 20 seconds with oxygen desaturation less than 80%. | Polygraphic recordings | Number of apnoeic episodes: 8 per day. Duration of episodes: 20-28 secs. | Polygraphic recordings | After about 10 s of central apnoea an abrupt decrease of the child's heart rate (more than 50% variation, from 160 bpm to 65 bpm) was recorded. The drop in the heart rate was followed by a new onset of thoracic efforts, associated with mixed central apnoea at the final stage, which lasted about 10 s, and then the episode resolved with a clear end-point. | The diagnostic tests and the results were not clearly described. The intervention and the post-intervention clinical condition were unclear. The adverse events were not identified. |
| Seppa-Moilanen et al. 2021<sup>40</sup> | 21 | Median: 35.7, inter-quartile range: 35.0 - 36.3 | Apnoea: central respiratory pauses longer than two breathing cycles and lasting 4 seconds or more. All obstructive breaths were noted regardless of the duration of apnoea.<br>AOP: an apnoea with a heart rate decrease to <100 beats/min, or a drop in oxygen saturation to under 80%, or it lasted longer than 20 s. | Nasal airflow by a pressure sensor, abdominal band. | Total of 3993 apnoeas in the recordings. | Monitor | For mixed apnoea, events accompanied by arousal were associated with longer durations and higher heart rates. In AOP, events with arousal had longer durations and lower heart rates. For mixed apnoea AOP, events with arousal exhibited longer durations but similar heart rates. | The details of the study subjects and the setting were unclear (study period was not stated). |

### 3.1 Relationship between apnoea duration and change in heart rate

A total of 31 articles investigated the change in heart rate compared with apnoea duration (Table 1), involving of 1008 neonates and 1734 recording sessions in total. Most studies (n = 26, 83.9%) measured heart rate using the electrocardiogram (ECG), while some used different monitors (Table 1). The relationship between heart rate changes and apnoea duration was examined using various methods, including correlation coefficients, linear regression models, and scatter plot diagrams. Most studies demonstrated that a longer apnoea duration is correlated with a greater reduction in heart rate. Six studies^4,24–28^ reported correlation coefficients, with all identifying a significant correlation between apnoea duration and metrics relating to decreased heart rate.

Five studies^29–33^ presented the mean change in heart rate during apnoea for different apnoea duration subgroups and were included in the meta-analysis. The average of the mean PMA of the neonates across the five studies was 32.84 ± 1.56 (mean ± SD). The pooled data across all five studies did not show a significant relationship between apnoea duration and heart rate change (Supplementary Figure 1, n = 5 studies; slope = −0.13, R^2^ = −0.02; r = −0.28, p = 0.10). However, data from Finer et al. (1992)^30^ showed an opposite trend to the other four studies, demonstrating a smaller change in heart rate for longer apnoeas. This might be due to the study’s unique definition of apnoea – the authors only recorded apnoeas associated with bradycardia or hypoxemia, excluding shorter pauses with less significant decrease in heart rate. Removing this study from further meta-analysis, the pooled data from the other four studies showed a negative relationship between apnoea duration and percentage change in heart rate during apnoea (Figure 2A, n = 4 studies; slope = −1.54, R^2^ = 0.36; r = −0.60, p = 0.0039).

**Figure 2:**
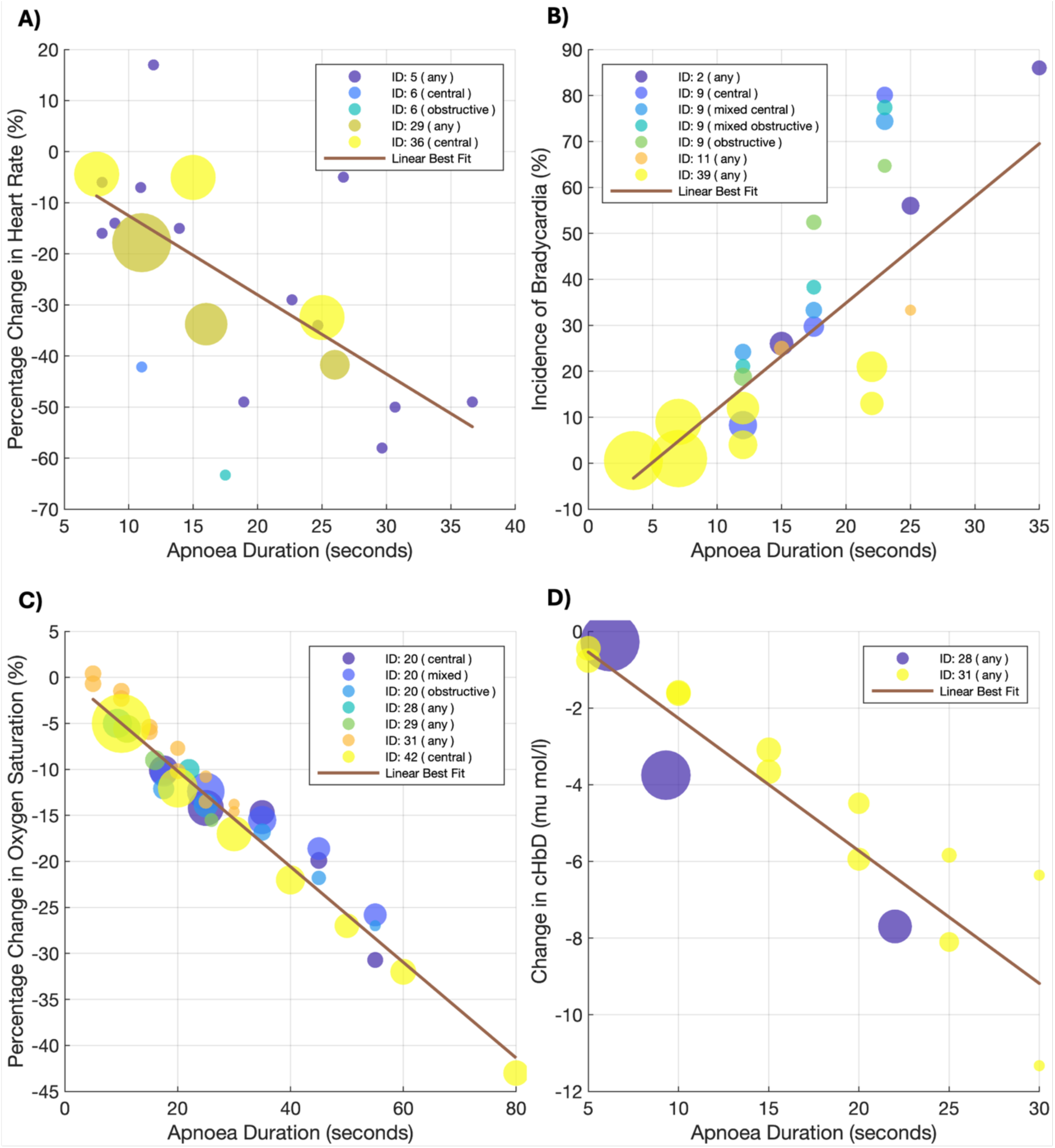
Meta-analysis of change in physiology compared with apnoea duration. (A) The percentage change in heart rate during apnoea compared with apnoea duration. (B) The percentage of apnoea events accompanied by bradycardia against apnoea duration. (C) The percentage change in oxygen saturation during apnoea compared with apnoea duration. (D) The change in cerebral haemoglobin difference (cHbD) during apnoea against apnoea duration. The size of the points indicates their weight in producing the linear best fit, calculated as the square root of the number of apnoea episodes associated with each point. ID: 2 – Gabriel et al. (1976)^17^, ID: 5 – Fenichel et al. (1980)^26^, ID: 6 – Vyas et al. (1981)^30^, ID: 9 - Henderson-Smart et al. (1986)^31^, ID: 11 – Mathew (1988)^32^, ID: 20 - Finer et al. (1992)^27^, ID: 28 – Urlesberger et al. (1999)^51^, ID: 29 – Carbone et al. (1999)^28^, ID: 31 – Pichler et al. (2003)^5^, ID: 36 – Beck et al. (2011)^29^, ID: 39– Marshall et al. (2019)^33^, ID: 42 – Varisco et al. (2022)^52^.

Five studies^19,30,34–36^ reported the percentage of different apnoea durations associated with a bradycardia event, defined as a decrease in heart rate to below 100 bpm; these were pooled in a separate meta-analysis. Gabriel et al. (1976) (b)^20^ also reported this relationship, but it was not included in the meta-analysis as the data and results were the same as Gabriel et al. (1976) (a)^19^. Most studies showed that longer apnoeas were associated with higher rates of bradycardia (Supplementary Figure 2, n = 5 studies; slope = 1.71, R^2^ = −0.05; r = 0.44, p = 0.0052). The study from Finer et al (1992)^30^ was again removed from the meta-analysis due to its unique definition of apnoea. The pooled data from the other four studies showed a positive correlation between apnoea duration and the percentage of apnoeas associated with bradycardia (Figure 2B, n = 4 studies, slope = 2.31, R^2^ = 0.57; r = 0.81, p < 0.0001).

The remaining studies (n = 12 studies) used a variety of heart rate metrics (e.g. absolute heart rate values during apnoea and multivariate linear analysis outcomes assessing the dependence of normalised percentage changes in heart rate on apnoea duration) and could not be pooled for additional meta-analysis due to limited data availability for each metric^21,37–47^ (Table 1, Supplementary Table 1). The majority (n = 11 studies) demonstrated a statistically significant relationship between change in heart rate and apnoea duration. In contrast, Curzi-Dascalova et al. (1989)^48^ studied 602 pauses in total, all lasting less than 12 seconds. Pauses of 10 to 12 seconds were rare (11 detected from all eligible babies) and did not induce higher cardiac deceleration than shorter pauses.

### 3.2 Relationship between apnoea duration and change in oxygen saturation

A total of 24 articles investigated the relationship between change in oxygen saturation with apnoea duration (Table 2), involving 1199 neonates and 1970 recording sessions in total. All the studies used a pulse oximeter to measure oxygen saturation.

**Table 2:** Summary of included studies reporting the relationship between apnoea duration and change in oxygen saturation. SD: standard deviation.

| Study | Sample size | Postmenstrual age at study (weeks) | Definition or inclusion criteria of apnoea/pause in breathing | Method of apnoea/pause measurement | Description of apnoea/pause in breathing | Method of outcome measurement | Main results | Quality assessment |
| --- | --- | --- | --- | --- | --- | --- | --- | --- |
| Muttitt et al. 1988<sup>21</sup> | 27 | Preterm. | See Table 1. | See Table 1. | See Table 1. | Pulse oximetry | A significant correlation was found between apnoea duration and decrease in oxygen saturation in the total population of infants with apnoea ($r = .388$, $P < .001$). Regression equation: percentage decrease in oxygen saturation = $3.778 + 0.338 * \text{duration of apnoea}$. | See Table 1. |
| Poets et al. 1991<sup>54</sup> | 31 (16 cases, 15 control) | Cases - mean: 40, range: 36 - 47.<br>Control - mean: 40, range: 35 - 45. | Apnoeic pauses: a cessation in breathing movements for at least 4 seconds.<br>Periodic breathing: a sequence of at least three apnoeic pauses, each separated by not more than 20 breaths. | Graseby pressure capsule, respitrace, thermistor. | Apnoea duration with episodes (s):<br>Cases:<br>periodic - median: 7.8, range: 6.2 - 19.2.<br>non-periodic - median: 11.9, range: 7.3 - 21.0.<br>Controls:<br>periodic - median: 5.4, range: 4.0 - 5.6.<br>non-periodic - median: 6.0, range: 4.0 - 8.5. | Pulse oximetry | The mean duration of apnoeic pauses associated with a hypoxaemic episode was significantly shorter during periodic breathing. | Unclear whether the two groups were comparable. Unclear whether the same criteria used for identification of cases and controls. Confounding factors were not identified. |
| Mathew et al. 1991<sup>37</sup> | 10 | Preterm. | See Table 1. | See Table 1. | See Table 1. | Pulse oximetry | Hypoxia or bradycardia developed during 24% of long apnoea ($\geq 15$ s) and only 7% of short apnoea ($< 15$ s). | See Table 1. |
| Upton et al. 1991<sup>4</sup> | 27 | Median: 32, range: 26 - 36. | See Table 1. | See Table 1. | See Table 1. | Pulse oximetry | There was a positive correlation between the reduction in oxygen saturation and the duration of apnoea. Reduction in oxygen saturation = $4.57 + 0.43 \times \text{duration of apnoea}$ ($r=0.41$, $p<0.0001$). | See Table 1. |
| Spear et al. 1992<sup>38</sup> | 26 | See Table 1. | See Table 1. | See Table 1. | See Table 1. | Pulse oximetry | Total number of desaturations < 90%, desaturations < 80% and < 90% in association with apnoea and/or bradycardia, longest duration of desaturation < 80%, and total duration of desaturation < 90% during sleep were all greater in the tracings with prolonged apnoea. | See Table 1. |
| Finer et al. 1992<sup>27</sup> | 47 | See Table 1. | See Table 1. | See Table 1. | See Table 1. | Pulse oximetry | As apnoea duration increased, the fall in oxygen saturation also increased ($r = 0.26$; $p = 0.0001$); this relationship was significant for all apnoea types and did not differ significantly between apnoea types or between treatment groups. There was a significant association between the pre-apnoea mean oxygen saturation and apnoea duration ($r = -0.14$; $p < 0.0001$). | See Table 1. |
| Abdulhamid et al. 1992<sup>55</sup> | 91 enrolled, 43 patients were <37 weeks gestation, and 25 of these were < 37 weeks postconceptional age at the time of evaluation. | Mean: 40.5, range: 34 - 88.<br>25 patients < 37. | central apnoea ≥ 20 seconds;<br>central apnoea < 20 seconds associated with bradycardia < 80 bpm for > 5 seconds in preterm infants . | Thoracic impedance, nasal / oral airflow. | A total of 184 episodes were recorded. | Pulse oximetry | The duration of obstructive apnoea ranged from 11 to 27 seconds, with 118 episodes (94%) associated with desaturation levels in the 71–84% range. Mixed apnoea episodes lasted between 10 and 45 seconds, with 25 episodes (47%) linked to desaturation levels in the 60–84% range. | The inclusion criteria and the population details were not stated. The details of the study subjects and the setting were unclear (study period was not stated). Confounding factors were not identified. Methods of statistical analysis were unclear. |
| Upton et al. 1992 (B)<sup>47</sup> | 24 | Median: 32, range: 26 - 36. | Apnoea: cessation of airflow for 5 seconds or longer. | Pneumotachograph, plethysmography, thoracic impedance. | 309 apnoeas. Episodes occurred in 60 of the 83 studies, with a median two apnoeas per study (range 0-19).<br>Mean apnoea duration recorded was 8.3 seconds (range 5-51 seconds). | Pulse oximetry | Positive correlation between oxygen saturation drops during apnoea and apnoea duration (oxygen saturation drop = (apnoea duration * 0.67) - 1.0, $r = 0.75$, $p < 0.0001$). | Unclear whether the participants were free of outcomes at the start of the study. |
| Poets et al. 1993<sup>42</sup> | 80 | Mean: 36.3, SD: 2.3. | See Table 1. | See Table 1. | See Table 1. | Pulse oximetry | The interval between onset of apnoeic pause and onset of desaturation has a median of 5.2 seconds, ranged between 0 and 15 seconds. | See Table 1. |
| Poets et al. 1995<sup>49</sup> | 96 | Median: 34, range: 29 - 37. | Apnoeic pause of 20 seconds or longer associated with desaturations were recorded. An association was considered present if the apnoeic pause began within 12 seconds of the desaturation. | Volume expansion capsule. | 23 desaturations in 7 infants were associated with prolonged apnoeic pauses ($\geq 20$ seconds). The duration of these pauses ranged from 21 to 120 seconds (median, 45 seconds). | Pulse oximetry | There was no significant correlation between the duration of the apnoeic pause and that of the desaturation ($r = 0.3$; $P > .05$, Spearman's rank correlation test). | The details of the study subjects and the setting were unclear (study period was not stated). The measurements of exposure were unclear. The strategies to deal with confounding factors were not stated. |
| Jenni et al. 1996<sup>24</sup> | 17 | Preterm | See Table 1. | See Table 1. | See Table 1. | Pulse oximetry | The duration of apnoea was correlated with the lowest oxygen saturation as follows: central apnoea: $r = -0.486$ ($p < 0.01$), obstructive apnoea: $r = -0.491$ ($p < 0.005$), and mixed apnoea: $r = -0.637$ ($p < 0.01$). | See Table 1. |
| Adams et al. 1997<sup>50</sup> | 30 | Mean: 32.8, SD: 2.3. | The duration of all respiratory pauses that led to a fall in oxygen saturation of $< 80\%$ for $> 4$ seconds. | Plethysmography, thermistor. | 156 pauses were recorded (40 apnoeas $> 15s$, 22 pauses with lower EELV (end-expiratory lung volume), 94 pauses with unchanged EELV) | Pulse oximetry | There was no significant correlation between the length of the respiratory pauses and the rate of desaturations ($r = -0.198$) | The inclusion criteria and the population details were not stated. Confounding factors were not identified. The method of statistical analysis was unclear. |
| Carbone et al. 1999<sup>28</sup> | 90 infants recruited, data were complete in 32 infants. | Mean: 36.9. | See Table 1. | See Table 1. | See Table 1. | Pulse oximetry | Longer apnoeas were significantly correlated with a lower saturation nadir, a longer duration of saturation $< 85\%$, a greater drop in saturation and a faster rate of saturation decline. | See Table 1. |
| Curzi-Dascalova et al. 2000<sup>41</sup> | 5 | Mean: 34, range: 33.6 - 34.7. | See Table 1. | See Table 1. | See Table 1. | Pulse oximetry | Linear mixed model analysis failed to demonstrate any significant dependence of the normalised changes in oxygen saturation on apnoea duration. | See Table 1. |
| Pichler et al. 2003<sup>5</sup> | See Table 1. | See Table 1. | See Table 1. | See Table 1. | See Table 1. | Pulse oximetry | Longer apnoea durations are associated with greater drops in arterial oxygen saturation in both the bradycardia and non-bradycardia groups. | See Table 1. |
| Nock et al. 2004<sup>39</sup> | 17 | Mean: 35, SD: 1. | See Table 1. | See Table 1. | See Table 1. | Pulse oximetry | Oxygen saturation during spontaneous apnoeas $\geq 15$ seconds fell to $86 \pm 9\%$, mean ± SD (range, 62%-99%). During apnoeic episodes $\geq 20$ seconds, saturation fell to $81 \pm 11\%$ (58%-92%). | See Table 1. |
| Cardot et al. 2007<sup>57</sup> | 36 | Premature | respiratory pause lasting $> 3$ seconds. | Thoracic impedance, pneumotachograph. | Mean apnoea duration was $5.5 \pm 1.3$ seconds. | Pulse oximetry | The duration of apnoeas with desaturation ($6.3 \pm 2.0$ seconds) was longer compared to the duration of apnoeas without desaturation ($5.2 \pm 1.0$ seconds). | The inclusion criteria and the population details were not stated. The details of the study subjects and the setting were unclear (study period was not stated). The measurements of exposure were unclear. The strategies to deal with confounding factors were not stated. |
| Tourneux et al. 2008<sup>56</sup> | 29 | Late preterm. | Central apnoeic events were defined as respiratory pauses lasting more than 3 seconds. | Plethysmography. | About 1900 apnoeic events were analysed. All lasted less than 20 s. | Pulse oximetry | Apnoeas with desaturation had longer durations compared to apnoeas without desaturation. | The inclusion criteria and the population details were not stated. The details of the study subjects and the setting were unclear (study period was not stated). The strategies to deal with confounding factors were not stated. |
| Elder et al. 2011<sup>48</sup> | Cases (n = 16).<br>Controls (n = 18). | Cases - median: 38, range: 35 - 45.<br>Controls - median: 37, range: 35 - 44. | Apnoea: cessation of nasal/oral airflow for $\geq 5$ seconds or a pause of $< 5$ seconds if there was a drop in oxygen saturation to $< 95\%$ or an associated 3% desaturation. | Thermistor. | Mean apnoea length (s): cases 6.9; controls 6.4. Longest apnoea (s): cases 17.4; controls 12.3. (Log-transformed, values expressed as geometric mean.) | Pulse oximetry | For all infants, there was a significant correlation between the lowest recorded oxygen saturation in active sleep for each infant and the longest apnoea duration recorded for each infant ($r = -0.530$, $p = 0.002$). This remained significant when group status was controlled for ($r = -0.436$, $p = 0.01$). | The groups were not comparable other than the presence of disease in cases or the absence of disease in controls. The measurements of exposure were unclear. Confounding factors were not identified. No statistical analysis performed. |
| Mohr et al. 2015<sup>25</sup> | 335 | Preterm | See Table 1. | See Table 1. | See Table 1. | Pulse oximeter | The area under the oxygen saturation curve increased approximately linearly with the duration of the apnoea (correlation coefficient: 0.88, p < 0.001). | See Table 1. |
| Fathabadi et al. 2017<sup>53</sup> | 20 | Preterm | Apnoea: a pause in respiration of 4 seconds or greater. | Respiratory motion. | Apnoea occurred concomitant with the hypoxic event in 555 (45 %) cases, including 495 (40 %) in which apnoea preceded the event and 165 events in which it occurred concurrently/subsequently (105 hypoxic events had both preceding and concurrent/subsequent apnoea). | Pulse oximetry | Longer apnoea durations were associated with longer hypoxia durations and a smaller oxygen saturation slope. | Confounding factors were not identified. Unclear whether the participants were free of the outcome at the start of the study. The measurements of outcomes were unclear. |
| Marshall et al. 2019<sup>33</sup> | 30 | Median: 30, inter-quartile range: 28-33. | See Table 1. | See Table 1. | See Table 1. | Pulse oximeter | In pooled data, the mean trajectories of oxygen saturation over the event window demonstrate more prominent instability after longer duration pauses, both for pauses that were isolated and clustered. The proportion of events followed by physiological instability increased with longer pause duration, and was higher for clustered events for all pause durations. | See Table 1. |
| Seppa-Moilanen et al. 2021<sup>40</sup> | 21 | Median: 35.7, inter-quartile range: 35.0 - 36.3 | See Table 1. | See Table 1. | See Table 1. | Pulse oxymeter | For central apnoea, events accompanied by arousal were associated with similar durations and higher oxygen saturation. For mixed apnoea, AOP and mixed apnoea AOP, events accompanied by arousal were associated with longer durations and higher oxygen saturation. | See Table 1. |
| Varisco et al. 2022<sup>52</sup> | 26 | Mean: 29.2. | CASEs: Count of central apnoea-suspected events. CASEs were then divided and grouped according to the length of their cessation of breathing L. The following lengths L were considered: 10s ≤ L < 20s, 20s ≤ L < 30s, 30s ≤ L < 40s, 40s ≤ L < 50s, 50s ≤ L < 60s, 60s ≤ L < 80s, L ≥ 80s. | Chest impedance. | A total number of 4756 CASEs were recorded. Number of CASEs for 10s ≤ L < 20s: 3140; 20s ≤ L < 30s: 655; 30s ≤ L < 40s: 404; 40s ≤ L < 50s: 202; 50s ≤ L < 60s: 106; 60s ≤ L < 80s: 127; L ≥ 80s: 122. | Pulse oxymeter | Longer apnoea durations were associated with greater decrease in oxygen saturation. The decrease was 5%, 12%, 17%, 22%, 27%, 32% and 43% for apnoea lengths of 10, 20, 30, 40, 50, 60 and 80 s, respectively. | The details of the study subjects and the setting were unclear (study period was not stated). The strategies to deal with confounding factors were not stated. No statistical analysis performed. |

As with the studies investigating changes in heart rate, a wide variety of oxygen saturation metrics and statistical analysis techniques were used. Eight studies^4,24,27,30,49–52^ reported correlations between apnoea duration and different oxygen saturation metrics. Six of these studies^4,24,27,30,49,50^ reported significant correlation coefficients, all indicating that longer apnoea durations were associated with greater decreases in oxygen saturation. Four of these significant correlations^4,24,30,49^ were based on the same oxygen saturation metric - the percentage decrease in oxygen saturation. The pooled correlation between these four correlation coefficients was 0.37 (n = 4 studies, p < 0.0001, 95% confidence interval: [0.34, 0.39]). In contrast, Poets et al. (1995)^51^ reported no significant correlation between the duration of the apnoeic pause and the duration of desaturation (r = 0.3; P > 0.05, Spearman’s rank correlation test). Similarly, Adams et al. (1997)^52^ found no significant correlation between the length of respiratory pauses and the rate of desaturations (r = −0.20).

Five studies^5,30,31,53,54^ presented the mean change in oxygen saturation during apnoea for different apnoea duration subgroups. The average of the mean PMA of participants in the five studies is 32.68 ± 2.48 weeks. Meta-analysis pooling the data from these studies showed a negative relationship between apnoea duration and change in oxygen saturation (Figure 2C, n = 5 studies, slope = −0.52, R^2^ = 0.95; r = − 0.98, p < 0.0001). Three studies^4,24,49^ used linear regression models to relate apnoea duration and change in oxygen saturation. The regression line slope from the pooled data (−0.51) were within the range of these models (Supplementary Figure 3, −0.34 for Muttitt et al. (1988)^24^, −0.43 for Upton et al. (1991)^4^, −0.67 for Upton et al. (1992)^49^).

The remaining studies used a variety of oxygen saturation metrics (e.g. area under the curve, lowest recorded oxygen saturation, and duration of apnoeas with and without desaturation), demonstrating significant correlations between apnoea duration and oxygen saturation. The data could not be pooled for additional meta-analysis due to limited data availability for each metric^21,28,36,40–42,44,55–59^ (Table 2, Supplementary Table 1).

### 3.3 Relationship between apnoea duration and change in cerebral oxygenation

Three studies examined the relationship between apnoea duration and cerebral oxygenation (Table 3). Changes in cerebral oxygenation were measured using Near Infra-Red Spectroscopy (NIRS). Jenni et al. (1996)^27^ reported the correlation coefficients between the amplitude of total haemoglobin concentration (tHb) and duration of apnoea as 0.55 for central apnoea, −0.005 for obstructive apnoea, and 0.7 for mixed apnoea. Urlesberger et al. (1999)^53^ and Pichler et al. (2003)^5^ presented the mean change in cerebral haemoglobin difference (cHbD) during apnoea for different apnoea duration groups. Meta-analysis of the two studies demonstrated that cHbD decreased more during apnoeas with longer durations (Figure 2D, n = 2 studies, slope = −0.35, R^2^ = 0.83; r = −0.92, p < 0.0001).

**Table 3:**
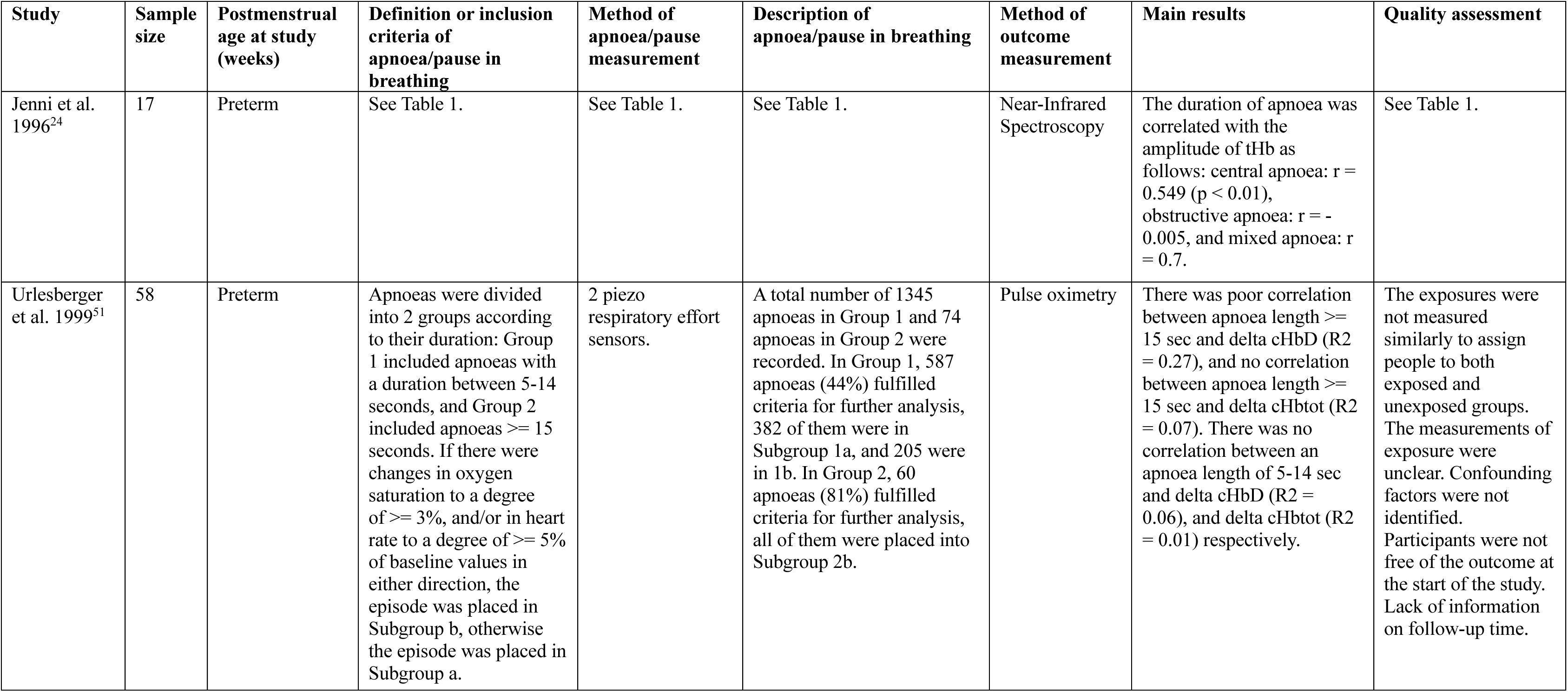
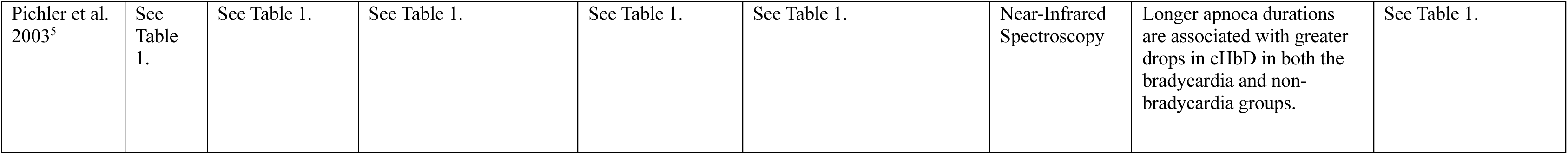
Summary of included studies reporting the relationship between apnoea duration and change in cerebral oxygenation. tHb: total haemoglobin concentration. cHbD: cerebral haemoglobin oxygenation index. cHbtot: concentration changes of total cerebral haemoglobin.

| Study | Sample size | Postmenstrual age at study (weeks) | Definition or inclusion criteria of apnoea/pause in breathing | Method of apnoea/pause measurement | Description of apnoea/pause in breathing | Method of outcome measurement | Main results | Quality assessment |
| --- | --- | --- | --- | --- | --- | --- | --- | --- |
| Jenni et al. 1996<sup>24</sup> | 17 | Preterm | See Table 1. | See Table 1. | See Table 1. | Near-Infrared Spectroscopy | The duration of apnoea was correlated with the amplitude of tHb as follows: central apnoea: $r = 0.549$ ($p < 0.01$), obstructive apnoea: $r = -0.005$, and mixed apnoea: $r = 0.7$. | See Table 1. |
| Urlesberger et al. 1999<sup>51</sup> | 58 | Preterm | Apnoeas were divided into 2 groups according to their duration: Group 1 included apnoeas with a duration between 5-14 seconds, and Group 2 included apnoeas $\geq 15$ seconds. If there were changes in oxygen saturation to a degree of $\geq 3\%$, and/or in heart rate to a degree of $\geq 5\%$ of baseline values in either direction, the episode was placed in Subgroup b, otherwise the episode was placed in Subgroup a. | 2 piezo respiratory effort sensors. | A total number of 1345 apnoeas in Group 1 and 74 apnoeas in Group 2 were recorded. In Group 1, 587 apnoeas (44%) fulfilled criteria for further analysis, 382 of them were in Subgroup 1a, and 205 were in 1b. In Group 2, 60 apnoeas (81%) fulfilled criteria for further analysis, all of them were placed into Subgroup 2b. | Pulse oximetry | There was poor correlation between apnoea length $\geq 15$ sec and delta cHbD ($R^2 = 0.27$), and no correlation between apnoea length $\geq 15$ sec and delta cHbtot ($R^2 = 0.07$). There was no correlation between an apnoea length of 5-14 sec and delta cHbD ($R^2 = 0.06$), and delta cHbtot ($R^2 = 0.01$) respectively. | The exposures were not measured similarly to assign people to both exposed and unexposed groups. The measurements of exposure were unclear. Confounding factors were not identified. Participants were not free of the outcome at the start of the study. Lack of information on follow-up time. |
| Pichler et al. 2003<sup>5</sup> | See Table 1. | See Table 1. | See Table 1. | See Table 1. | See Table 1. | Near-Infrared Spectroscopy | Longer apnoea durations are associated with greater drops in cHbD in both the bradycardia and non-bradycardia groups. | See Table 1. |

**Table 4:** Summary of included studies reporting the relationship between apnoea duration and change in cerebral blood volume.

| Study | Sample size | Postmenstrual age at study (weeks) | Definition or inclusion criteria of apnoea/pause in breathing | Method of apnoea/pause measurement | Description of apnoea/pause in breathing | Method of outcome measurement | Main results | Quality assessment |
| --- | --- | --- | --- | --- | --- | --- | --- | --- |
| Pichler et al. 2003<sup>5</sup> | See Table 1. | See Table 1. | See Table 1. | See Table 1. | See Table 1. | Near-Infrared Spectroscopy | Longer apnoea durations are associated with greater drops in cerebral blood volume (CBV) in the bradycardia group, but not in the non-bradycardia group. | See Table 1. |

### 3.4 Relationship between apnoea duration and change in cerebral blood volume

Only one study from Pichler et al. (2003)^5^ reported the relationship between apnoea duration and cerebral blood volume (Table 4). The changes in cerebral blood volume were measured using NIRS. The study population comprised two subgroups: a bradycardia group (bradycardia was defined as a heart rate decrease to below 80 beats per minute, n = 20) and a non-bradycardia group (n = 19). Episodes of apnoea with associated bradycardia were matched with episodes of apnoea without bradycardia based on the duration and PMA of the neonates. The cerebral blood volume decreased significantly with increased apnoea duration in the bradycardia group, but the relationship was not significant in the non-bradycardia group.

### 3.5 Factors which modulate the physiological response to apnoea

While most studies showed that longer apnoea durations correspond to greater physiological changes, there is considerable variation, particularly in heart rate changes (Figure 2). Studies included in this review have investigated whether apnoea type and the use of theophylline can affect the relationship between apnoea duration and changes in physiology. Using a meta-analysis, we also explored whether PMA modulates the relationship.

#### 3.5.1 Apnoea type

Several studies^24,25,27^ showed that central and obstructive apnoeas could lead to different physiological responses. Muttitt et al. (1988)^24^ identified a significant correlation between apnoea duration and heart rate decrease in central apnoeas (computer-diagnosed: r = 0.19, p < 0.0001; nurse-diagnosed: r = 0.20, p < 0.0001), but not in obstructive or mixed apnoea groups. Suichies et al. (1989)^25^ reported significant positive correlations between the percentage of apnoeas associated with bradycardias and apnoea duration in central apnoeas (r = 0.65, p < 0.01) and mixed apnoeas (r = 56, p < 0.05), but the correlation was not significant for obstructive apnoeas. Similarly, Jenni et al. (1996)^27^ found that the minimum heart rate during apnoea was negatively correlated with apnoea duration in central apnoeas (r = −0.77, p < 0.01) and mixed apnoeas (r = −0.69, p < 0.01) only, but again, no significant correlation was observed in obstructive apnoeas. However, with regard to oxygen saturation, Jenni et al. (1996)^27^ observed a significant correlation between apnoea duration and oxygen saturation for all apnoea types (central: r = −0.49, p < 0.01; obstructive: r = −0.49, p < 0.05; mixed: r = −0.64, p < 0.01).

Studies shown in Figure 2A included four data points associated with central apnoea, one data point associated with obstructive apnoea, and 16 data points involving any apnoea type. Meta-analysis on apnoea type was not performed for these studies due to limited data availability. The meta-analysis of the studies presented in Figures 2B and 2C indicated that apnoea type played a significant role in modulating the relationship between bradycardia incidence rate and apnoea duration (Supplementary Table 2), as well as between changes in oxygen saturation and apnoea duration (Supplementary Table 3).

#### 3.5.2 Theophylline

There is limited evidence for theophylline as a modulating factor. Muttitt et al. (1988) ^24^ found in a sub-group analysis that the correlation between apnoea duration and decrease in heart rate was significant in a theophylline-treated group (r = 0.16, p < 0.0001) but not in the untreated group. Theophylline did not significantly affect the decrease in oxygen saturation in this study. However, the correlation between apnoea duration and decrease in oxygen saturation was the strongest in the theophylline-treated group (r = 0.43, p < 0.0001). Similarly, Upton et al. (1991)^4^ found that theophylline did not reduce the slope of the reduction in oxygen saturation for the duration of the apnoeic attack (treated: r = 0.45, p < 0.0001; untreated: r = 0.24, p < 0.0001).

#### 3.5.3 Postmenstrual age

While none of the included studies analysed the effect of age on neonates’ responses to apnoea, the meta-analysis of the studies shown in Figure 2C revealed that PMA significantly modulated the relationship between changes in oxygen saturation and apnoea duration (Supplementary Table 4). A post hoc analysis showed that for neonates with a PMA of 30, 32, 34, and 36 weeks, the predicted apnoea durations associated with a drop in oxygen saturation greater than 10% were 18.9, 20.0, 21.5, and 23.4 seconds, respectively (i.e. younger neonates have a significant drop in oxygen saturation with shorter apnoeas). In the mixed-effects model analysis comparing percentage change in heart rate with apnoea duration, PMA was not a significant factor (Supplementary Table 5). PMA was not incorporated into the other mixed-effects models due to a lack of information.

### 3.6 Quality assessment

A summary of the outcomes of the quality assessments can be found in Tables 1-4, and the detailed results from the JBI checklists are provided in Supplementary Tables 6-9. None of the papers satisfied all the requirements in the checklists. The results of overall risk of bias assessments can be found in Supplementary Table 10. Five studies (12.2%) had a low overall risk of bias, 21 studies (51.2%) had a moderate overall risk of bias, and 15 studies (36.6%) had a high overall risk of bias. Among the 15 studies included in the meta-analyses in Figure 2 and the combined correlation factors in Section 3.2, 7 had a high risk of bias, 6 had a moderate risk, and 2 had a low risk.

## 4. Discussion

This systematic review and meta-analysis investigated how the duration of apnoea, or pauses in breathing, correlates with changes in physiology in preterm neonates. Overall, the meta-analysis demonstrated that the minimum value and the extent of decrease in heart rate, oxygen saturation, and cerebral oxygenation during apnoea were significantly correlated with the duration of apnoea. This aligns with the outcomes of most studies included in this review. In contrast, a change in cerebral blood volume was only correlated with apnoea duration in the group of neonates with bradycardia, but not in the non-bradycardia group. However, this was only investigated in one study, and therefore the results require further validation.

Although there is a strong relationship between changes in physiology and apnoea duration, there is nevertheless considerable variation (Figure 2), with the R^2^ values indicating that changes in heart rate exhibited greater variation than changes in oxygen saturation. In the meta-analysis, changes in physiological parameters were assessed as the percentage change during apnoea compared to a baseline period before the apnoea. As neonates generally have a baseline oxygen saturation above 90%, and clinical teams intervene if saturation decreases below 90% for a significant period, the differences in baselines across studies were minimal. In contrast, baseline heart rate can vary much more significantly between individual neonates, which might partly explain this difference in variation between the models. Moreover, differences between studies, such as in measurement techniques or ventilatory support may account for variation.

In addition to variation across studies, there is considerable variation across individual apnoeas, as observed in the included papers, such as Upton et al. (1991)^4^. Significant and recurrent physiological fluctuations from apnoea may have clinical implications both acutely and long-term, with studies suggesting associations with increased incidence of ROP^7^ and cognitive deficits later in life^8,9^. Understanding the factors which modulate the relationship between apnoea duration and changes in physiology could allow for the identification of neonates that are at particular risk from large changes in physiology following even short apnoeas, and conversely those neonates who remain physiologically stable. Apnoea type, the use of theophylline and the neonate’s PMA were identified as significant factors modulating the relationship between apnoea duration and changes in physiology. Through meta-analysis, we demonstrate that PMA significantly modulates the relationship between apnoea duration and oxygen saturation. This result suggests that different durations of breathing pause should be used as apnoea alarm limits for neonates with different PMA to reduce oxygen desaturations. In particular, we propose that for infants younger than 32 weeks PMA, an apnoea alarm limit of 15 seconds is considered. Investigation of other factors modulating the relationship was limited by the number of studies included in this review and the availability of data for each outcome examined. Insufficient data prevented analysis of factors such as pathological conditions, ventilation modes and medications other than theophylline. Future studies are warranted to identify other modulating factors.

This study was limited by the small sample sizes of most included studies, which may introduce potential bias in the results. Additionally, the publication years of the studies were relatively old, with more than half published over 30 years ago. The methods of measurement and clinical techniques likely differ significantly from those used today. Only five papers showed a low overall risk of bias, while the remaining studies had either moderate or high risks of bias. Many papers had missing or unclear information on important topics. For example, less than half (40.9%) reported the sex of the neonates. There was significant inconsistency between papers in the definitions of apnoea, bradycardia, and other events. Furthermore, useful information was sometimes presented only in figures, requiring us to extract data points from those figures, which could lead to inaccuracies. Nevertheless, meta-analysis demonstrated consistent results across studies and provide a basis for future work investigating the relationship between apnoea duration and change in physiology.

## 5. Conclusion

This systematic review and meta-analysis provides strong evidence that longer apnoea durations directly correlate with greater deterioration in cardiorespiratory and cerebral parameters in preterm neonates. Most critically, we identified postmenstrual age as a key determinant of physiological vulnerability – with younger infants more likely to experience oxygen desaturation from shorter apnoeas. Current standard monitor settings using 20-second thresholds for all preterm infants are likely inadequate for the most vulnerable babies. We recommend clinical implementation of PMA-stratified apnoea alarm thresholds, with a threshold of 15 seconds for infants <32 weeks PMA. Additionally, central apnoeas warrant closer monitoring than obstructive events due to their stronger association with physiological deterioration. These findings are readily translatable to clinical practice and should inform newborn care unit monitoring protocols to prevent potentially harmful physiological instability in our most vulnerable patients.

## Author Contributions

YC contributed to the study design, screened the search results, extracted data, completed the quality assessment and drafted the initial manuscript. CH designed the study, provided supervision and screened the search results. CZ contributed to screening the search results and quality assessment. LBa, OF, VK, RP and ZS contributed to screening the search results. MH developed the search strategy. FU, LBe and MV provided mentorship and supervision. All authors critically reviewed and revised the manuscript.

## Consent statement

Patient consent was not required.

## Codes availability

Code for the meta-analysis can be found at: https://github.com/Yiru-C/apnoea_physiology_meta_analysis.git

## Data availability

All extracted data are provided in Supplementary Table 1.

## Data Availability

All extracted data are provided in Supplementary Table 1.

