## Supplementary Material for "Relationship between apnoea duration and changes in physiology in preterm neonates: a systematic review and meta-analysis"

### Supplementary Methods

#### MEDLINE database via Ovid platform

Medline (Ovid MEDLINE® Epub Ahead of Print, In-Process & Other Non-Indexed Citations, Ovid MEDLINE® Daily and Ovid MEDLINE®) 1946 to present

1 infant/ or infant, newborn/ or infant, low birth weight/ or infant, small for gestational age/ or infant, very low birth weight/ or infant, extremely low birth weight/ or infant, postmature/ or infant, premature/ or infant, extremely premature/ or (infant* or newborn* or new-born* or perinatal* or neonatal* or neo-natal* or postnatal* or post-natal* or baby or babies or neonate* or neo-nate*).ti,ab,kf.

2 apnea/ or breath holding/ or (apno* or apne* or breath hold* or periodic breath* or pause in breath* or cessation of breath* or AOP or respiratory pause* or intermittent breath* or intermittent respiration or inter breath interval* or stop breath*).ti,ab,kf.

3 heart rate/ or bradycardia/ or tachycardia/ or oxygen saturation/ or hypoxia/ or cerebral blood volume/ or brain ischemia/ or hypoxia-ischemia, brain/ or (heart rate* or cardiac rate* or heartbeat* or pulse rate* or bradycardia or tachycardia or oxygen saturation or desat* or oxygen deprivation or lack of oxygen or oxygen starvation or hypoxia or hypoxemia or hypoxeamia or SpO2 or SaO2 or PaO2 or cerebral blood volume or CBV or cerebral blood flow or brain ischemia or cerebral ischeamia or brain blood flow or brain blood supply or brain hemodynamics or brain haemodynamics or brain perfusion or cerebral oxygenation).ti,ab,kf.

4 1 and 2 and 3

5 exp animals/ not humans/

6 4 not 5

**Embase database via Ovid platform**

Embase 1974 to present

1 infant/ or baby/ or high risk infant/ or hospitalized infant/ or newborn/ or prematurity/ or (infant* or newborn* or new-born* or perinatal* or neonatal* or neo-natal* or postnatal* or post-natal* or baby or babies or neonate* or neo-nate*).ti,ab,kf

2 *apnea/ or *apnea attack/ or *newborn apnea/ or *newborn apnea attack/ or *breath holding/ or (apno* or apne* or "breath hold*" or "periodic breath*" or "pause in breath*" or "cessation of breath*" or AOP or "respiratory pause*" or "intermittent breath*" or "intermittent respiration" or "inter breath interval*" or "stop breath*").ti,ab,kf.

3 heart rate/ or bradycardia/ or tachycardia/ or oxygen saturation/ or hypoxia/ or hypoxemia/ or newborn hypoxia/ or brain blood volume/ or brain ischemia/ or ("heart rate*" or "cardiac rate*" or heartbeat* or "pulse rate*" or bradycardia or tachycardia or "oxygen saturation" or desat* or "oxygen deprivation" or "lack of oxygen" or "oxygen starvation" or hypoxia or hypoxemia or hypoxeamia or SpO2 or SaO2 or PaO2 or "cerebral blood volume" or CBV or "cerebral blood flow" or "brain ischemia" or "cerebral ischeamia" or "brain blood flow" or "brain blood supply" or "brain hemodynamics" or "brain haemodynamics" or "brain perfusion" or "cerebral oxygenation").ti,ab,kf.

4 1 and 2 and 3

5 exp animal/ not human/

6 4 not 5

7 Systematic review.ti,ab. not (trial or study).ti.

8 (review.ab. and review.pt.) not trial.ti.

9 we searched.ab. and (review.ti. or review.pt.)

10 update review.ab.

11 (databases adj4 searched).ab.

12 (conference abstract or conference paper or "conference review" or editorial).pt.

13 7 or 8 or 9 or 10 or 11 or 12

14 6 not 13

**PsycINFO database via Ovid platform**

PsycINFO 1806 to present

1 premature birth/ or (infant* or newborn* or new-born* or perinatal* or neonatal* or neo-natal* or postnatal* or post-natal* or baby or babies or neonate* or neo-nate*).ti,ab,id.

2 apnea/ or (apno* or apne* or "breath hold*" or "periodic breath*" or "pause in breath*" or "cessation of breath*" or AOP or "respiratory pause*" or "intermittent breath*" or "intermittent respiration" or "inter breath interval*" or "stop breath*").ti,ab,id.

3 heart rate/ or bradycardia/ or tachycardia/ or anoxia/ or cerebral blood flow/ or cerebral ischemia/ or ("heart rate*" or "cardiac rate*" or heartbeat* or "pulse rate*" or bradycardia or tachycardia or "oxygen saturation" or desat* or "oxygen deprivation" or "lack of oxygen" or "oxygen starvation" or hypoxia or hypoxemia or hypoxeamia or SpO2 or SaO2

or PaO2 or "cerebral blood volume" or CBV or "cerebral blood flow" or "brain ischemia" or "cerebral ischeamia" or "brain blood flow" or "brain blood supply" or "brain hemodynamics" or "brain haemodynamics" or "brain perfusion" or "cerebral oxygenation").ti,ab,id.

4 1 and 2 and 3

**Cochrane Library platform**

#1 [mh ^infant] OR [mh ^"infant, newborn"] OR [mh ^"infant, low birth weight"] OR [mh ^"infant, small for gestational age"] OR [mh ^"infant, very low birth weight"] OR [mh ^"infant, extremely low birth weight"] OR [mh ^"infant, postmature"] OR [mh ^"infant, premature"] OR [mh ^"infant, extremely premature"] OR (infant*:ti,ab,kw OR newborn*:ti,ab,kw OR new-born*:ti,ab,kw OR perinatal*:ti,ab,kw OR neonatal*:ti,ab,kw OR neo-natal*:ti,ab,kw OR postnatal*:ti,ab,kw OR post-natal*:ti,ab,kw OR baby:ti,ab,kw OR babies:ti,ab,kw OR neonate*:ti,ab,kw OR neo-nate*:ti,ab,kw)

#2 [mh ^apnea] OR [mh ^"breath holding"] OR (apno*:ti,ab,kw OR apne*:ti,ab,kw OR ("breath" NEXT hold*):ti,ab,kw OR ("periodic" NEXT breath*):ti,ab,kw OR ("pause in" NEXT breath*):ti,ab,kw OR ("cessation of" NEXT breath*):ti,ab,kw OR AOP:ti,ab,kw OR ("respiratory" NEXT pause*):ti,ab,kw OR ("intermittent" NEXT breath*):ti,ab,kw OR "intermittent respiration":ti,ab,kw OR ("inter breath" NEXT interval*):ti,ab,kw OR ("stop" NEXT breath*):ti,ab,kw)

#3 [mh ^"heart rate"] OR [mh ^bradycardia] OR [mh ^tachycardia] OR [mh ^"oxygen saturation"] OR [mh ^hypoxia] OR [mh ^"cerebral blood volume"] OR [mh ^"brain ischemia"] OR [mh ^"hypoxia-ischemia, brain"] OR (("heart" NEXT rate*):ti,ab,kw OR ("cardiac" NEXT rate*):ti,ab,kw OR heartbeat*:ti,ab,kw OR ("pulse" NEXT rate*):ti,ab,kw OR bradycardia:ti,ab,kw OR tachycardia:ti,ab,kw OR "oxygen saturation":ti,ab,kw OR desat*:ti,ab,kw OR "oxygen deprivation":ti,ab,kw OR "lack of oxygen":ti,ab,kw OR "oxygen starvation":ti,ab,kw OR hypoxia:ti,ab,kw OR hypoxemia:ti,ab,kw OR hypoxeamia:ti,ab,kw OR SpO2:ti,ab,kw OR SaO2:ti,ab,kw OR PaO2:ti,ab,kw OR "cerebral blood volume":ti,ab,kw OR CBV:ti,ab,kw OR "cerebral blood flow":ti,ab,kw OR "brain ischemia":ti,ab,kw OR "cerebral ischeamia":ti,ab,kw OR "brain blood flow":ti,ab,kw OR "brain blood supply":ti,ab,kw OR "brain hemodynamics":ti,ab,kw OR "brain haemodynamics":ti,ab,kw OR "brain perfusion":ti,ab,kw OR "cerebral oxygenation":ti,ab,kw)

#4 #1 and #2 and #3

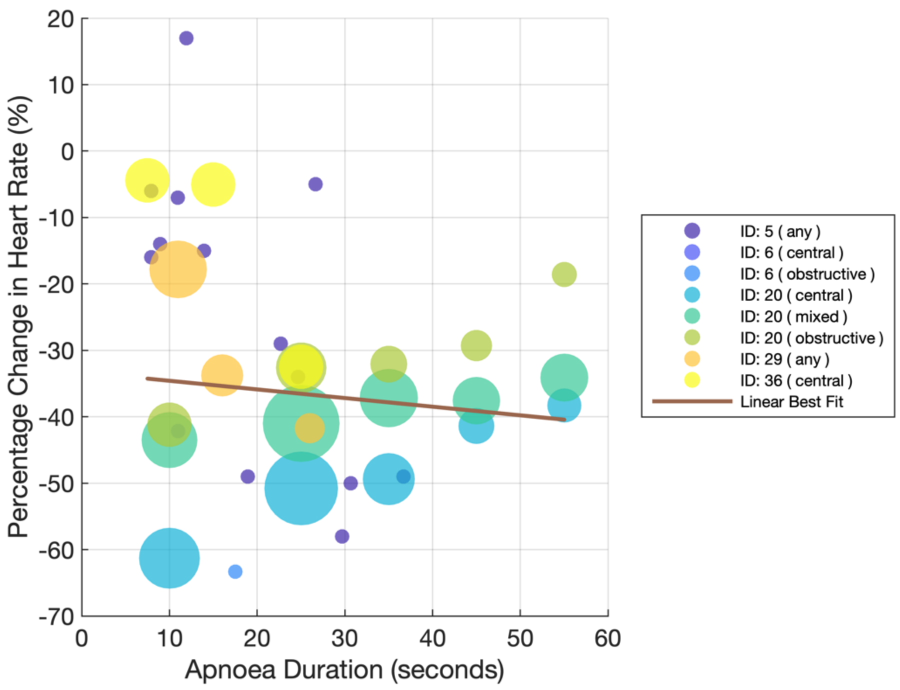


**Supplementary Figure 1**: The percentage change in heart rate during apnoea on y-axis against apnoea duration on the x-axis. This plot includes the data from Finer et al. (1992). The size of the points indicates their weight in producing the linear best fit, calculated as the square root of the number of apnoea episodes associated with each point. ID: 5 – Fenichel et al. (1980)[26], ID: 6 – Vyas et al. (1981)[30], ID: 20 – Finer et al. (1992)[27], ID: 29 – Carbone et al. (1999)[28], ID: 36 – Beck et al. (2011)[29].

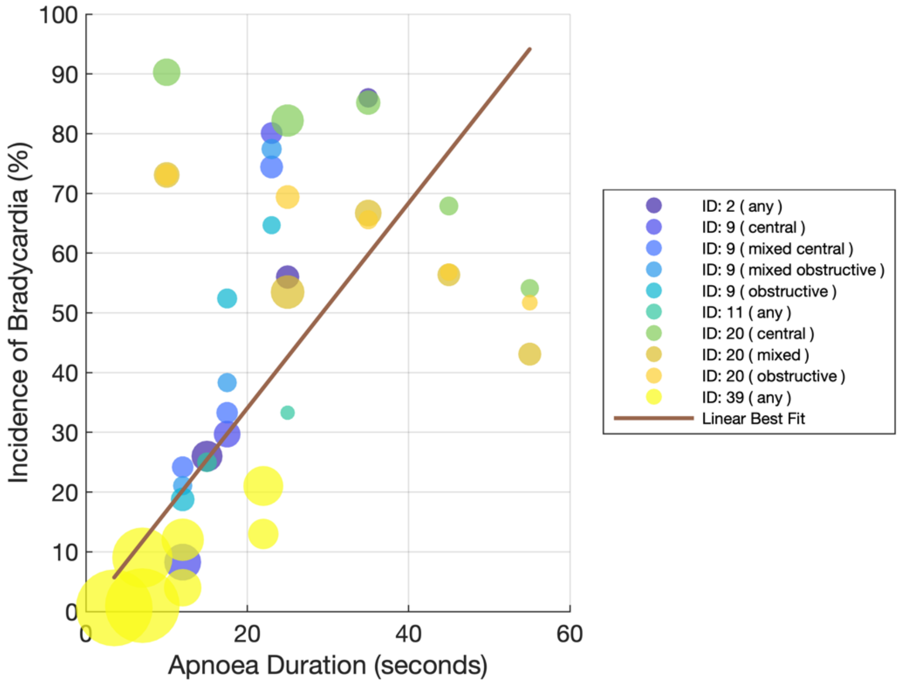


**Supplementary Figure 2**: The plot displays percentage of apnoea events accompanied by bradycardia on the y-axis against apnoea duration on the x-axis. This plot includes the data from Finer et al. (1992). The size of the points indicates their weight in producing the linear best fit, calculated as the square root of the number of apnoea episodes associated with each point. ID: 2 – Gabriel et al. (1976)[17], ID: 9 - Henderson-Smart et al. (1986)[31], ID: 11 – Mathew (1988)[32], ID: 20 - Finer et al. (1992)[27], ID: 39 – Marshall et al. (2019)[33].

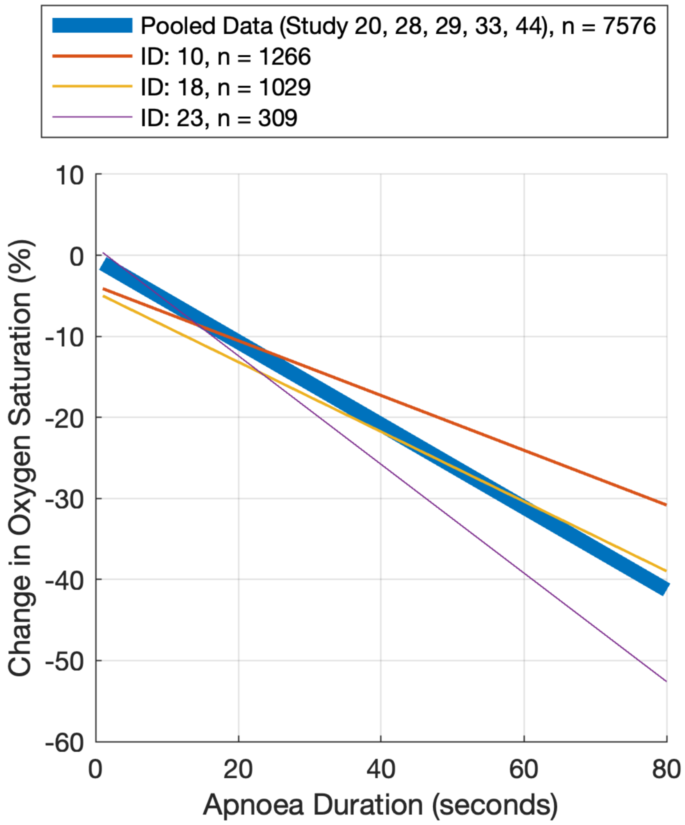


**Supplementary Figure 3**: This plot displays the change in oxygen saturation on y-axis versus apnoea duration on x-axis. The lines are the best-fit linear regression lines. The three thin lines (red, yellow, purple) represent the regression lines reported by individual studies. The thick blue line represents the regression lines of the data pooled from Gabriel et al. (1976)[17], Henderson-Smart et al. (1986)[31], Mathew (1988)[32], and Marshall et al. (2019)[33]. The number n and the thickness of the lines represents the number of apnoea events associated with each line. ID: 10 – Muttitt et al. (1988)[21], ID: 18 – Upton et al. (1991)[4], ID: 23 – Upton et al. (1992)[47].

**Supplementary Table 1:** Full data set extracted from all the papers included in the review. See the separate excel file. ECG: Electrocardiogram. SD: standard deviation. $SaO_2$: oxygen saturation. tHb: total haemoglobin concentration. cHbD: cerebral haemoglobin oxygenation index. cHbtot: concentration changes of total cerebral haemoglobin.

**Supplementary Table 2: Results of the statistical analysis investigating if apnoea type modulates the relationship between bradycardia incidence and apnoea duration.** Fixed effects coefficients from the linear mixed-effects model with the formula: "bradycardia incidence ~ 1 + ID + apnoea type * apnoea duration". * indicates $p < 0.05$, ** indicates $p < 0.01$, *** indicates $p < 0.001$.

| **Name** | **Estimate** | **Standard Error** | **t Statistic** | **Degree of Freedom** | **p Value** |
|---|---|---|---|---|---|
| (Intercept) | -7.28 | 2.77 | -2.63 | 14 | 0.020* |
| Apnoea type (central) | -58.6 | 19.6 | -2.99 | 14 | 0.010** |
| Apnoea_type (obstructive) | -24.6 | 29.8 | -0.83 | 14 | 0.42 |
| Apnoea_type (mixed central) | -29.6 | 29.2 | -1.02 | 14 | 0.33 |
| Apnoea_type (mixed obstructive) | -37.4 | 34.6 | -1.08 | 14 | 0.30 |
| Apnoea_duration | 1.66 | 0.241 | 6.88 | 14 | < 0.0001*** |
| Apnoea_type (central):Apnoea_duration | 4.34 | 1.25 | 3.46 | 14 | 0.0038** |
| Apnoea_type (obstructive):Apnoea_duration | 2.75 | 1.80 | 1.53 | 14 | 0.15 |
| Apnoea_type (mixed central):Apnoea_duration | 2.98 | 1.60 | 1.87 | 14 | 0.08 |
| Apnoea_type (mixed obstructive):Apnoea_duration | 3.51 | 1.89 | 1.86 | 14 | 0.08 |

**Supplementary Table 3: Results of the statistical analysis investigating if apnoea type modulates the relationship between oxygen saturation and apnoea duration.** Fixed effects coefficients from the linear mixed-effects model with the formula: "oxygen saturation fall ~ 1 + ID + apnoea type * apnoea duration". * indicates $p < 0.05$, ** indicates $p < 0.01$, *** indicates $p < 0.001$.

| **Name** | **Estimate** | **Standard Error** | **t Statistic** | **Degree of Freedom** | **pValue** |
|---|---|---|---|---|---|
| (Intercept) | -0.20 | 0.52 | -0.38 | 31 | 0.71 |
| Apnoea type (mixed) | -2.56 | 1.50 | -1.71 | 31 | 0.10 |
| Apnoea_type (obstructive) | -4.49 | 2.12 | -2.12 | 31 | 0.042* |
| Apnoea_type (any) | 1.89 | 1.10 | 1.71 | 31 | 0.097 |
| Apnoea_duration | -0.53 | 0.01 | -35.6 | 31 | < 0.0001*** |
| Apnoea_type (mixed):Apnoea_duration | 0.14 | 0.04 | 3.32 | 31 | 0.0023** |
| Apnoea_type (obstructive):Apnoea_duration | 0.15 | 0.07 | 2.31 | 31 | 0.028* |
| Apnoea_type (any):Apnoea_duration | -0.03 | 0.06 | -0.52 | 31 | 0.60 |

**Supplementary Table 4: Results of the statistical analysis investigating if postmenstrual age (PMA) and apnoea type modulates the relationship between oxygen saturation and apnoea duration.** Fixed effects coefficients from the linear mixed-effects model with the formula: "oxygen saturation fall ~ 1 + ID + apnoea type * apnoea duration + PMA * apnoea duration". * indicates $p < 0.05$, ** indicates $p < 0.01$, *** indicates $p < 0.001$.

| **Name** | **Estimate** | **Standard Error** | **t Statistic** | **Degree of Freedom** | **pValue** |
|---|---|---|---|---|---|
| (Intercept) | 41.1 | 12.3 | 3.34 | 29 | 0.0023** |
| PMA | -1.41 | 0.41 | -3.37 | 29 | 0.0021** |
| Apnoea type (mixed) | 0.52 | 1.57 | 0.33 | 29 | 0.74 |
| Apnoea_type (obstructive) | -1.41 | 2.03 | -0.69 | 29 | 0.49 |
| Apnoea_type (any) | 10.5 | 2.72 | 3.87 | 29 | 0.0006*** |
| Apnoea_duration | -2.13 | 0.44 | -4.83 | 29 | < 0.0001*** |
| PMA:Apnoea_duration | 0.05 | 0.015 | 3.63 | 29 | 0.0011** |
| Apnoea_type (mixed):Apnoea_duration | 0.03 | 0.05 | 0.55 | 29 | 0.59 |
| Apnoea_type (obstructive):Apnoea_duration | 0.03 | 0.06 | 0.53 | 29 | 0.60 |
| Apnoea_type (any):Apnoea_duration | -0.36 | 0.10 | -3.45 | 29 | 0.0017** |

**Supplementary Table 5: Results of the statistical analysis investigating if postmenstrual age (PMA) modulates the relationship between heart rate drops and apnoea duration.** Fixed effects coefficients from the linear mixed-effects model with the formula: "heart rate fall ~ 1 + ID + apnoea type * apnoea duration + PMA * apnoea duration". * indicates $p < 0.05$, ** indicates $p < 0.01$, *** indicates $p < 0.001$.

| **Name** | **Estimate** | **Standard Error** | **t Statistic** | **Degree of Freedom** | **pValue** |
|---|---|---|---|---|---|
| (Intercept) | 93.8 | 98.7 | 0.95 | 17 | 0.36 |
| PMA | -2.61 | 2.91 | -0.90 | 17 | 0.38 |
| Apnoea_duration | -1.18 | 6.03 | -0.20 | 17 | 0.85 |
| PMA:Apnoea_duration | -0.013 | 0.18 | -0.08 | 17 | 0.94 |

**Supplementary Table 6(A):** JBI Critical Appraisal Checklist for the included cross-sectional studies (Part A).

| **Variables assessed** | **Daily et al. (1969)** | **Fenichel et al. (1980)** | **Werthammer et al. (1983)** | **Muttitt et al. (1988)** | **Curzi-Dascalova et al. (1989)** | **Suichies et al. (1989)** | **Hodgman et al. (1990)** | **Mathew et al. (1991)** | **Abdulhamid et al. (1992)** | **Finer et al. (1992)** | **Poets et al. (1993)** | **Poets et al. (1995)** |
|---|---|---|---|---|---|---|---|---|---|---|---|---|
| Were the criteria for inclusion in the sample clearly defined? | No | No | No | No | No | No | Yes | No | No | No | No | Yes |
| Were the study subjects and the setting described in detail? | Yes | No | No | Yes | Unclear | Unclear | Yes | Yes | Unclear | Unclear | Unclear | Unclear |
| Was the exposure measured in a valid and reliable way? | Yes | Unclear | Yes | Yes | Unclear | Unclear | Unclear | Yes | Yes | Unclear | Unclear | Unclear |
| Were objective, standard criteria used for measurement of the condition? | Yes | Yes | Yes | Yes | Yes | Yes | Yes | Yes | Yes | Yes | Yes | Yes |
| Were confounding factors identified? | No | No | No | Yes | Yes | No | No | No | No | No | No | Yes |
| Were strategies to deal with confounding factors stated? | N/A | N/A | N/A | Yes | Yes | N/A | N/A | N/A | N/A | N/A | N/A | No |
| Were the outcomes measured in a valid and reliable way? | Yes | No | Yes | Unclear | Yes | Yes | Yes | Yes | Yes | Yes | Yes | Yes |
| Was appropriate statistical analysis used? | N/A | N/A | N/A | Yes | N/A | Yes | Unclear | N/A | Unclear | Yes | N/A | Yes |

**Supplementary Table 6(B):** JBI Critical Appraisal Checklist for the included cross-sectional studies (Part B).

| Variables assessed | Jenni et al. (1996) | Adams et al. (1997) | Carbone et al. (1999) | Curzi-Dascalova et al. (2000) | Nock et al. (2004) | Cardot et al. (2007) | Tourneux et al. (2008) | Beck et.al (2011) | Mohr et al. (2015) | Seppa-Moilanen et al. (2021) | Varisco et al. (2022) |
|---|---|---|---|---|---|---|---|---|---|---|---|
| Were the criteria for inclusion in the sample clearly defined? | No | No | No | Yes | Yes | No | No | Yes | No | Yes | Yes |
| Were the study subjects and the setting described in detail? | Yes | Yes | No | Unclear | Unclear | Unclear | Unclear | Unclear | Yes | Unclear | Unclear |
| Was the exposure measured in a valid and reliable way? | Unclear | Yes | Unclear | Unclear | Unclear | Unclear | Yes | Yes | Yes | Yes | Yes |
| Were objective, standard criteria used for measurement of the condition? | Yes | Yes | Yes | Yes | Yes | Yes | Yes | Yes | Yes | Yes | Yes |
| Were confounding factors identified? | No | No | No | Yes | No | Yes | No | Yes | No | Yes | Yes |
| Were strategies to deal with confounding factors stated? | N/A | N/A | N/A | Yes | N/A | No | N/A | No | N/A | Yes | No |
| Were the outcomes measured in a valid and reliable way? | Yes | Yes | Yes | Yes | Yes | Yes | Yes | Yes | Yes | Yes | Yes |
| Was appropriate statistical analysis used? | Yes | Unclear | Unclear | Yes | Yes | Yes | Yes | Yes | Yes | Yes | N/A |

**Supplementary Table 7:** JBI Critical Appraisal Checklist for the included cohort studies.

| **Variables assessed** | **Gabriel et al. (1976)** | **Storrs (1977)** | **Vyas et al. (1981)** | **Southall et al. (1983)** | **Henderson-Smart et al. (1986)** | **Mathew (1988)** | **Waggener et al. (1989)** | **Upton et al. (1991)** | **Spear et al. (1992)** | **Upton et al. (1992) (A)** | **Upton et al. (1992) (B)** | **Urlesberger et al. (1999)** | **Pichler et al. (2003)** | **Fathabadi et al. (2017)** | **Marshall et al. (2019)** |
|---|---|---|---|---|---|---|---|---|---|---|---|---|---|---|---|
| Were the two groups similar and recruited from the same population? | N/A | N/A | N/A | Yes | N/A | N/A | N/A | N/A | No | N/A | N/A | Yes | Unclear | N/A | N/A |
| Were the exposures measured similarly to assign people to both exposed and unexposed groups? | N/A | N/A | N/A | Yes | N/A | N/A | N/A | N/A | Yes | N/A | N/A | No | Yes | N/A | N/A |
| Was the exposure measured in a valid and reliable way? | Unclear | Unclear | Yes | Yes | Yes | Yes | Yes | Unclear | Yes | Unclear | Yes | Unclear | Yes | Yes | Yes |
| Were confounding factors identified? | No | No | Yes | No | Yes | No | No | Yes | Yes | Yes | Yes | No | No | No | Yes |
| Were strategies to deal with confounding factors stated? | N/A | No | Yes | N/A | No | N/A | N/A | No | Yes | Yes | Yes | N/A | N/A | N/A | Yes |
| Were the groups/participants free of the outcome at the start of the study (or at the moment of exposure)? | Unclear | Unclear | No | No | Unclear | Yes | Unclear | Yes | Unclear | Unclear | Unclear | No | Unclear | Unclear | Unclear |
| Were the outcomes measured in a valid and reliable way? | Unclear | Yes | Yes | Yes | Yes | Yes | Yes | Yes | Yes | Unclear | Yes | Yes | Yes | No | Yes |
| Was the follow up time reported and sufficient to be long enough | Unclear | Unclear | Unclear | Unclear | Unclear | Yes | Unclear | Unclear | Unclear | Unclear | Unclear | Unclear | Unclear | Unclear | Unclear |

| | | | | | | | | | | | | | | | |
|---|---|---|---|---|---|---|---|---|---|---|---|---|---|---|---|
| for outcomes to occur? | | | | | | | | | | | | | | | |
| Was follow up complete, and if not, were the reasons to loss to follow up described and explored? | Unclear | Unclear | Unclear | Unclear | Unclear | Yes | Unclear | Unclear | Unclear | N/A | Unclear | Unclear | Unclear | Unclear | Unclear |
| Were strategies to address incomplete follow up utilized? | No | No | No | No | No | No | No | No | No | N/A | No | No | No | No | No |
| Was appropriate statistical analysis used? | N/A | Unclear | N/A | N/A | Yes | Yes | Yes | Yes | No | Yes | Yes | Yes | Yes | Yes | Yes |

**Supplementary Table 8:** JBI Critical Appraisal Checklist for the included case control studies.

| **Variables assessed** | **Poets & Southall (1991)** | **Elder et al. (2011)** |
|---|---|---|
| Were the groups comparable other than the presence of disease in cases or the absence of disease in controls? | Unclear | No |
| Were cases and controls matched appropriately? | Yes | Yes |
| Were the same criteria used for identification of cases and controls? | Unclear | Yes |
| Was exposure measured in a standard, valid and reliable way? | Yes | Unclear |
| Was exposure measured in the same way for cases and controls? | Yes | Yes |
| Were confounding factors identified? | No | No |
| Were strategies to deal with confounding factors stated? | N/A | N/A |
| Were outcomes assessed in a standard, valid and reliable way for cases and controls? | Yes | Yes |
| Was the exposure period of interest long enough to be meaningful? | Yes | Yes |
| Was appropriate statistical analysis used? | Yes | N/A |

**Supplementary Table 9:** JBI Critical Appraisal Checklist for the included case report study.

| **Variables assessed** | **Falsaperla et al. (2019)** |
|---|---|
| Were patient's demographic characteristics clearly described? | Yes |
| Was the patient's history clearly described and presented as a timeline? | Yes |
| Was the current clinical condition of the patient on presentation clearly described? | Yes |
| Were diagnostic tests or assessment methods and the results clearly described? | No |
| Was the intervention(s) or treatment procedure(s) clearly described? | Unclear |
| Was the post-intervention clinical condition clearly described? | Unclear |
| Were adverse events (harms) or unanticipated events identified and described? | No |
| Does the case report provide takeaway lessons? | Yes |

**Supplementary Table 10:** Overall risk of bias assessment.

| Study | Checklist | Internal Validity 'No' Responses | Overall Risk of Bias |
|---|---|---|---|
| Gabriel et al. (1976) | Cohort | 2 | High |
| Storrs (1977) | Cohort | 3 | High |
| Vyas et al. (1981) | Cohort | 2 | High |
| Southall et al. (1983) | Cohort | 3 | High |
| Henderson-Smart et al. (1986) | Cohort | 2 | High |
| Mathew (1988) | Cohort | 2 | High |
| Waggener et al. (1989) | Cohort | 2 | High |
| Upton et al. (1991) | Cohort | 2 | High |
| Spear et al. (1992) | Cohort | 2 | High |
| Upton et al. (1992) (A) | Cohort | 0 | Low |
| Upton et al. (1992) (B) | Cohort | 1 | Moderate |
| Urlesberger et al. (1999) | Cohort | 4 | High |
| Pichler et al. (2003) | Cohort | 2 | High |
| Fathabadi et al. (2017) | Cohort | 3 | High |
| Marshall et al. (2019) | Cohort | 1 | Moderate |
| Poets & Southall (1991) | Case-Control | 1 | Moderate |
| Elder et al. (2011) | Case-Control | 2 | High |
| Daily et al. (1969) | Cross-Sectional | 1 | Moderate |
| Fenichel et al. (1980) | Cross-Sectional | 2 | High |
| Werthammer et al. (1983) | Cross-Sectional | 1 | Moderate |
| Muttitt et al. (1988) | Cross-Sectional | 0 | Low |
| Curzi-Dascalova et al. (1989) | Cross-Sectional | 0 | Low |
| Suichies et al. (1989) | Cross-Sectional | 1 | Moderate |
| Hodgman et al. (1990) | Cross-Sectional | 1 | Moderate |
| Mathew et al. (1991) | Cross-Sectional | 1 | Moderate |
| Abdulhamid et al. (1992) | Cross-Sectional | 1 | Moderate |
| Finer et al. (1992) | Cross-Sectional | 1 | Moderate |
| Poets et al. (1993) | Cross-Sectional | 1 | Moderate |
| Poets et al. (1995) | Cross-Sectional | 1 | Moderate |

| | | | |
|---|---|---|---|
| Jenni et al. (1996) | Cross-Sectional | 1 | Moderate |
| Adams et al. (1997) | Cross-Sectional | 1 | Moderate |
| Carbone et al. (1999) | Cross-Sectional | 1 | Moderate |
| Curzi-Dascalova et al. (2000) | Cross-Sectional | 0 | Low |
| Nock et al. (2004) | Cross-Sectional | 1 | Moderate |
| Cardot et al. (2007) | Cross-Sectional | 1 | Moderate |
| Tourneux et al. (2008) | Cross-Sectional | 1 | Moderate |
| Beck et.al (2011) | Cross-Sectional | 1 | Moderate |
| Mohr et al. (2015) | Cross-Sectional | 1 | Moderate |
| Seppa-Moilanen et al. (2021) | Cross-Sectional | 0 | Low |
| Varisco et al. (2022) | Cross-Sectional | 1 | Moderate |
| Falsaperla et al. (2019) | Case Report | 2 | High |
